# Interim report: Safety and immunogenicity of an inactivated vaccine against SARS-CoV-2 in healthy chilean adults in a phase 3 clinical trial

**DOI:** 10.1101/2021.03.31.21254494

**Authors:** Susan M Bueno, Katia Abarca, Pablo A González, Nicolás MS Gálvez, Jorge A Soto, Luisa F Duarte, Bárbara M Schultz, Gaspar A Pacheco, Liliana A González, Yaneisi Vázquez, Mariana Ríos, Felipe Melo-González, Daniela Rivera-Pérez, Carolina Iturriaga, Marcela Urzúa, Angélica Dominguez, Catalina A Andrade, Roslye V Berrios, Gisela Canedo-Marroquín, Camila Covián, Daniela Moreno-Tapia, Farides Saavedra, Omar P Vallejos, Paulina Donato, Pilar Espinoza, Daniela Fuentes, Marcela González, Paula Guzmán, Paula Muñoz-Venturelli, Carlos M Pérez, Marcela Potin, Alvaro Rojas, Rodrigo Fasce, Jorge Fernández, Judith Mora, Eugenio Ramírez, Aracelly Gaete-Argel, Aarón Oyarzún-Arrau, Fernando Valiente-Echeverría, Ricardo Soto-Rifo, Daniela Weiskopf, Alessandro Sette, Gang Zeng, Weining Meng, José V González-Aramundiz, Alexis M Kalergis

## Abstract

**Background:** The ongoing COVID-19 pandemic has had a significant impact worldwide, with an incommensurable social and economic burden. The rapid development of safe and protective vaccines against this disease is a global priority. CoronaVac is a vaccine prototype based on inactivated SARS-CoV-2, which has shown promising safety and immunogenicity profiles in pre-clinical studies and phase 1/2 trials in China. To this day, four phase 3 clinical trials are ongoing with CoronaVac in Brazil, Indonesia, Turkey, and Chile. This article reports the safety and immunogenicity results obtained in a subgroup of participants aged 18 years and older enrolled in the phase 3 Clinical Trial held in Chile.

**Methods:** This is a multicenter phase 3 clinical trial. Healthcare workers aged 18 years and older were randomly assigned to receive two doses of CoronaVac or placebo separated by two weeks (0-14). We report preliminary safety results obtained for a subset of 434 participants, and antibody and cell-mediated immunity results obtained in a subset of participants assigned to the immunogenicity arm. The primary and secondary aims of the study include the evaluation of safety parameters and immunogenicity against SARS-CoV-2 after immunization, respectively. This trial is registered at clinicaltrials.gov (NCT04651790).

**Findings:** The recruitment of participants occurred between November 27^th^, 2020, until January 9^th^, 2021. 434 participants were enrolled, 397 were 18-59 years old, and 37 were ≥60 years old. Of these, 270 were immunized with CoronaVac, and the remaining 164 participants were inoculated with the corresponding placebo. The primary adverse reaction was pain at the injection site, with a higher incidence in the vaccine arm (55.6%) than in the placebo arm (40.0%). Moreover, the incidence of pain at the injection site in the 18-59 years old group was 58.4% as compared to 32.0% in the ≥60 years old group. The seroconversion rate for specific anti-S1-RBD IgG was 47.8% for the 18-59 years old group 14 days post immunization (p.i.) and 95.6% 28 and 42 days p.i. For the ≥60 years old group, the seroconversion rate was 18.1%, 100%, and 87.5% at 14, 28, and 42 days p.i., respectively. Importantly, we observed a 95.7% seroconversion rate in neutralizing antibodies for the 18-59 years old group 28 and 42 days p.i. The ≥60 years old group exhibited seroconversion rates of 90.0% and 100% at 28 and 42 days p.i. Interestingly, we did not observe a significant seroconversion rate of anti-N-SARS-CoV-2 IgG for the 18-59 years old group. For the participants ≥60 years old, a modest rate of seroconversion at 42 days p.i. was observed (37.5%). We observed a significant induction of a T cell response characterized by the secretion of IFN-γ upon stimulation with Mega Pools of peptides derived from SARS-CoV-2 proteins. No significant differences between the two age groups were observed for cell-mediated immunity.

**Interpretation:** Immunization with CoronaVac in a 0-14 schedule in adults of 18 years and older in the Chilean population is safe and induces specific IgG production against the S1-RBD with neutralizing capacity, as well as the activation of T cells secreting IFN-γ, upon recognition of SARS-CoV-2 antigens.

**Funding:** Ministry of Health of the Chilean Government; Confederation of Production and Commerce, Chile; Consortium of Universities for Vaccines and Therapies against COVID-19, Chile; Millennium Institute on Immunology and Immunotherapy.

## Introduction

Severe acute respiratory syndrome coronavirus 2 (SARS-CoV-2) is the emerging pathogen responsible for coronavirus disease 2019 (COVID-19) ^1–3^. This virus was first described in December 2019 in Wuhan, China, and it is the source of the ongoing pandemic, which has resulted by March 2021 in almost 130 million cases and over 2,7 million deaths worldwide ^4^. Due to its novelty and impact on human health, international efforts are focused on generating vaccines to counteract COVID-19. Epidemiological studies show that individuals aged ≥60 and those with chronic conditions are more susceptible to severe disease, frequently resulting in death ^5,6^. To date, more than 180 vaccines are under development, with over 80 of them in clinical trials, 18 undergoing phase 3 or phase 4 clinical trials, and 10 approved for emergency use ^7^. While many different vaccine platforms are being used and explored, most of them rely on a single viral component, namely the full-length Spike (S) protein or the receptor-binding domain (RBD) of Spike^7,8^.

CoronaVac is an inactivated SARS-CoV-2 vaccine, developed by Sinovac Life Sciences Co., Ltd. (Beijing, China) ^9^. Sinovac received approval from the NMPA in China in April 2020 to conduct phase 1/2 studies of this vaccine candidate in that country after demonstrating that it was safe and immunogenic in different animal models, such as rodents and non-human primates ^9^. Phase 1/2 clinical trials in China were carried out as randomized, double-blind, and placebo-controlled studies. In these trials, two vaccination schedules were evaluated: two doses separated by 14 days (0-14) or 28 days (0-28) ^10,11^. Overall, these studies reported one severe adverse event (SAE) possibly related to the vaccine during the phase 2 trial with participants aged 18-59, which resolved within three days ^10^. There were eight SAEs in the ≥60 years old age group, but all were unrelated to the vaccine ^11^. Both phase 2 trials showed that this vaccine induces neutralizing antibodies 14 days after the second dose ^10,11^. The neutralizing antibody seroconversion rate was above 92% in participants aged 18-59, and above 94% for participants aged ≥60, using two 3 µg doses in the 0-28 schedule ^10,11^. The results from these clinical trials suggest that this vaccine is safe and likely induces a protective immune response against SARS-CoV-2.

Currently, four phase 3 clinical trials are evaluating the efficacy of CoronaVac, and are being carried out in Brazil, Turkey, Indonesia, and Chile. Here, we report an interim analysis of safety and immunogenicity parameters upon immunization of the Chilean population with CoronaVac or placebo in healthcare workers aged 18-59 and ≥60 in a 0-14 days vaccination schedule. A total of 434 participants were evaluated, 397 aged 18-59 and 37 aged ≥60. Given that this vaccine carries multiple SARS-CoV-2 antigens, the characterization of the humoral and cellular immune response was extended to other components of the viral proteome beyond Spike. Taken together, this is the first report characterizing the cellular and humoral immune responses elicited by CoronaVac in a population other than the Chinese. Our results indicate that CoronaVac is safe and immunogenic in adults aged 18-59 and ≥60 in the Chilean population.

## Materials and Methods

### Study design, randomization, and participants

This clinical trial (clinicaltrials.gov NCT04651790) was conducted in Chile at eight different sites. The study protocol was conducted according to the current Tripartite Guidelines for Good Clinical Practices, the Declaration of Helsinki ^12^, and local regulations. The trial protocol was reviewed and approved by the Institutional Scientific Ethical Committee of Health Sciences, Pontificia Universidad Católica de Chile, Approval #200708006 (Committee members: Claudia Uribe PUC/Committee president; Colomba Cofré/Committee Vice-president; Andréa Villagrán/Executive secretary; Jorge Muñoz/External Lawyer; Gustavo Kaltwasser/External member; Alysa Garay/Community representative; Marisa Torres/Public Health Department; Carolina Méndez/Speech therapy representative; Luis Villarroel/Public Health Department; Pablo Brockman/Respiratory Diseases in Children Department. Website: http://eticayseguridad.uc.cl/comite-etico-cientifico-facultad-de-medicina-uc.html). Trial execution was approved by the Chilean Public Health Institute (#24204/20). Written informed consent was obtained from each participant before enrollment. The study included healthcare workers who were in contact with possible or confirmed cases of COVID-19 ≥18 years old. Participants were inoculated with either two doses of 3 µg (600SU) of CoronaVac or placebo at 0- and 14-days post the first immunization (p.i.). Exclusion criteria included, among others, history of confirmed symptomatic SARS-CoV-2 infection, pregnancy, allergy to vaccine components, and immunocompromised conditions. Well-controlled medical conditions were allowed. A complete list of inclusion/exclusion criteria is provided in the study protocol.

Participants were randomly assigned to immunization with CoronaVac or injection with placebo in a 1:1 ratio. A subgroup of participants was assigned to the immunogenicity arm, and they received randomly either CoronaVac or placebo (3:1 ratio). The randomization process was done using a sealed enveloped system integrated into the electronic Case Report Forms (eCRF) in the OpenClinica platform. All participants, blind investigators, and laboratory staff were masked to arm allocation. A total of 434 participants were enrolled up to January 9^th^, 2021. The immunogenicity arm includes 190 participants. Table 1 summarizes the characteristics of the participants, and Figure 1 shows the study profile.

**Table 1.** Characteristics of the Participants at Baseline.

| Characteristic | 18-59 years old<br>(n = 397) | ≥ 60 years old<br>(n = 37) | Total<br>(n = 434) | p-value |
| --- | --- | --- | --- | --- |
| **Age mean, SD** | 38.2 ± 9.7 | 64.0 ± 4.3 | 40.4 ± 11.8 |  |
| **Inoculation** |  |  |  |  |
| Vaccine; n (%) | 245 (61.7) | 25 (67.6) | 270 (62.2) | 0.482 |
| Placebo; n (%) | 152 (38.3) | 12 (32.4) | 164 (37.8) |  |
| **Sex** |  |  |  |  |
| Women; n (%) | 251 (63.2) | 17 (45.9) | 268 (61.8) | 0.039 |
| Men; n (%) | 146 (36.8) | 20 (54.1) | 166 (38.2) |  |
| **Ethnicity** |  |  |  |  |
| White; n (%) | 370 (93.2) | 37 (100.0) | 407 (93.8) | 0.152 |
| Other; n (%) | 27 (6.8) | 0 (0.0) | 27 (6.2) |  |

P-values are for comparison between total numbers in each characteristic.

**Figure 1.**
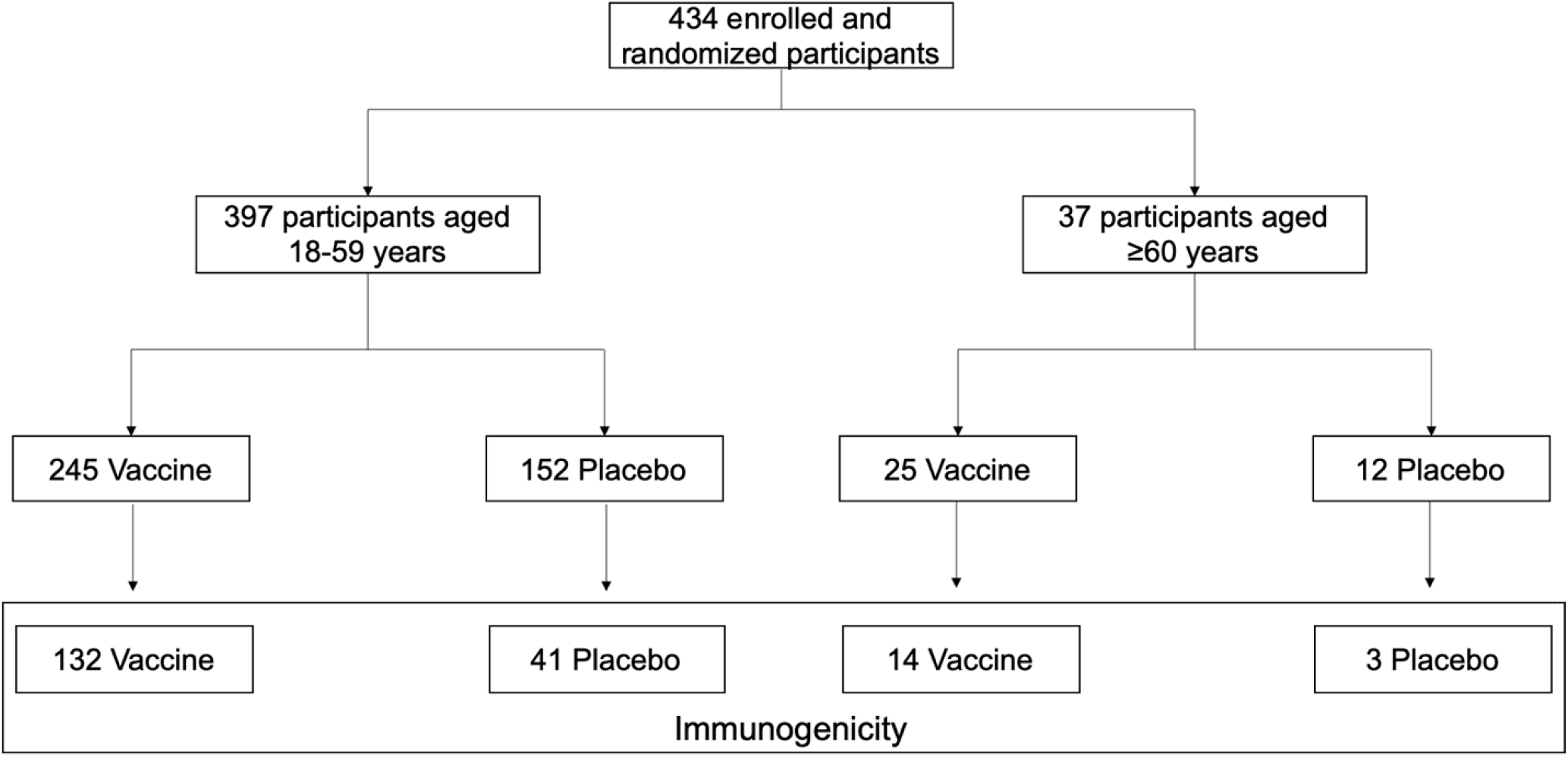
Study profile. Recruitment of volunteers as of January 08^th^, 2021.

### Procedures

CoronaVac consists of 3 µg of β-propiolactone inactivated SARS-CoV-2 (strain CZ02) with aluminum hydroxide as an adjuvant in 0.5 mL ^9^. More details on excipients and the Placebo can be found in the Supplementary Material. A non-blind study nurse was in charge of administering intramuscularly in the deltoid area the contents of ready-to-use syringes loaded with either CoronaVac or placebo to participants. For the immunogenicity arm, blood samples were obtained at different time points and were used for the isolation of sera and PBMC. For sera samples, 20 mL of blood were collected. For PBMC isolation, 30 mL of blood were collected in heparinized tubes. Details regarding isolation of sera and PBMCs can be found in the Supplementary Material.

To assess the presence of anti-SARS-CoV-2 antibodies, blood samples from 39 participants obtained at days 0 (baseline), 14, 28, and 42 p.i. were analyzed. The quantitative measurement of human IgG antibodies against the RBD domain of the S1 protein (S1-RBD) and against the N protein of SARS-CoV-2 was determined using the RayBio COVID-19 (SARS-CoV-2) Human Antibody Detection Kit (Indirect ELISA method) (Cat #IEQ-CoVS1RBD-IgG & #IEQ-CovN-IgG). Details on the characteristics and the methodology of these kits can be found on the Supplementary Material. The neutralizing capacities of the antibodies in the samples of the participants were evaluated through the SARS-CoV-2 surrogate virus neutralization test (sVNT) kit from Genscript (cat. number L00847-A). Assays were performed according to the instructions of the manufacturer and further details can be found on the Supplementary Material. To assess the cellular immune response, ELISPOT and flow cytometry assays were performed using PBMCs from 36 participants. Stimulus included in these assays considers the use of Mega Pools (MPs) of peptides derived from SARS-CoV-2 proteins, previously described ^13^. Two MPs composed of peptides from the S protein (MP-S) and the remaining proteins of the viral particle (MP-R) were used. Also, two MPs composed of peptides from the whole proteome of SARS-CoV-2 (CD8-A and CD8-B) were used. Corresponding positives and negative controls were held. A total of 3×10^5^ cells in 50 µL of media were added to each well containing 50 µL of media with the corresponding stimulus. For ELISPOT assays, cells were incubated for 48h at 37°C, 5% CO_2_. Further details regarding the ELISPOT protocol can be found in the Supplementary Materials. To characterize T cells and the production of cytokines by these populations, flow cytometry assays were performed. A total of 3×10^5^ cells per well were stimulated with the same stimulus indicated above. 24h after incubation, samples were stained to evaluate the expression of surface and intracellular markers. Further details on the antibodies used and the protocol can be found in the Supplementary Materials.

### Outcomes

The primary aim was to evaluate the frequency of solicited and unsolicited AEs that occur during 7 days after each dose, stratified by age group (aged 18-59 and ≥60). Grading for solicited and unsolicited AEs can be found in detail in the Table S1, S2, and S3 of the Supplementary Materials. Secondary safety endpoints include the frequency of any other AE occurring 28 days after each dose, SAEs, and Events of Special Interest in any moment, and their relationship with the investigational vaccine. Secondary immunogenic endpoints considered, among others, the assessment of the presence of anti-SARS-CoV-2 antibodies and the cellular immune response elicited by the vaccine in a subgroup of participants, at days 0, 28, and 42 p.i.. A complete list of outcomes can be found in the study protocol.

### Statistical analysis

To determine the sample size to use in this trial, the parameters used were a protection rate of the vaccine of (µ= 0.5); an incidence rate in the population group of 6%; α = 0.05; 1-β = 0.9. A total of 1,068 participants need to be recruited in each group. Considering a 10% rate of withdrawal, the sample size in each group was defined as 1,175.

Safety analysis considers all AEs (solicited and unsolicited) that occurred after each injection. All AEs were coded and grouped according to MedDRA (Medical Dictionary for Regulatory Activities) methodology. Solicited AEs (local and systemic) are presented in a summarized form according to frequency, their maximum intensity, and duration per participant. Rates were compared between several groups (vaccine v/s placebo; 18-59 v/s ≥60 years old; first v/s second dose) using a two-tailed Pearson’s Chi-square or Fisher’s exact test. Unsolicited AEs with a frequency of 1% or more; SAEs; and Events of Special Interest are presented.

To evaluate statistical differences in the immunogenicity date, one- or two-way non-parametric ANOVAs (Friedman test) with the corresponding *post hoc* test (Dunn’s test corrected for multiple comparisons) or Wilcoxon tests or two-tailed Student’s t-test (for comparisons between two groups) were performed depending on the assay. The significance level was set at 0.05. All data were analyzed with GraphPad Prism 9.0.1. or SPSS 17.0.

## Results

### 1. CoronaVac shows a favorable safety profile in adults aged 18-59 and ≥60 in the Chilean population

Participants of this phase of the study were recruited between November 27^th^, 2020, and January 9^th^, 2021. 434 participants were enrolled, with 397 aged 18-59 and 37 aged 60-75. 268 participants were women (61.7%), and 166 were men (38.3%). The mean age (SD) was 39.7 (± 11.8) years in the placebo arm and 40.9 (± 11.9) in the vaccine arm. 319 participants received two doses of either CoronaVac or placebo (286 in the 18-59 age group and 33 in the ≥60 age group). The vaccination schedule for both groups was 0-14. The demographic characteristics of the participants are summarized in Table 1.

A list of local and systemic solicited AEs observed after each vaccine dose is shown in Table 2. The most-reported solicited local AEs was pain at the injection site, with an incidence of 55.6% in the vaccine arm as compared to 40.0% in the placebo arm (p-value = 0.015). Symptoms usually started after vaccination and were resolved within two days, with predominantly mild (grade 1) severity (98%, 91%, and 100%, respectively). Local AEs were more frequent in the vaccine arm (278 events) than in the placebo arm (78 events). Headaches were the most common solicited systemic AEs with a frequency of 48.5% in the vaccine arm and 48.8% in the placebo arm. Intensity of AEs was mostly mild or moderate. No serious adverse events (SAEs) nor Events of Special Interest were reported in either arm. Significant differences were observed between age groups regarding the frequency of local and systemic AEs (Table S4): pain was present in 47.1% of the 18-59 years old group in contrast to 29.7% in the ≥60 years old group (p-value = 0.009), while headache was present in 42.4% and 27.0%, respectively (p-value = 0.026). This association remained present in the vaccine arm but not in the placebo arm.

**Table 2.** **Solicited Local and Systemic Adverse Events after inoculation in Participants, classified by arm and age group**

| Adverse Reaction | First dose<br>(n =434) |  |  | Second dose<br>(n =319) |  |  | Both Doses<br>(n =319) |  |  |
| --- | --- | --- | --- | --- | --- | --- | --- | --- | --- |
|  | Placebo<br>(n=164) | Vaccine<br>(n=270) | p-value | Placebo<br>(n=80) | Vaccine<br>(n=239) | p-value | Placebo<br>(n=80) | Vaccine<br>(n=239) | p-value |
| **Local reactions** |  |  |  |  |  |  |  |  |  |
| Pain; n (%) | 39 (23.8) | 117 (43.3) | <0.001 | 16 (20.0) | 73 (30.5) | 0.069 | 32 (40.0) | 133 (55.6) | 0.015 |
| <60 years | 37 (24.3) | 113 (46.1) | <0.001 | 15 (20.8) | 68 (31.8) | 0.077 | 30 (41.7) | 125 (58.4) | 0.014 |
| ≥60 years | 2 (16.7) | 4 (16.0) | 0.999 | 1 (12.5) | 5 (20.0) | 0.999 | 2 (25.0) | 8 (32.0) | 0.999 |
| Induration (%) | 1 (0.6) | 8 (3.0) | 0.163 | 0 (0.0) | 15 (6.3) | 0.015 | 0 (0.0) | 21 (8.8) | 0.006 |
| <60 years | 1 (0.7) | 7 (2.9) | 0.161 | 0 (0.0) | 13 (6.1) | 0.043 | 0 (0.0) | 18 (8.4) | 0.009 |
| ≥60 years | 0 (0.0) | 1 (4.0) | 0.999 | 0 (0.0) | 2 (8.0) | 0.999 | 0 (0.0) | 3 (12.0) | 0.56 |
| Pruritus (%) | 4 (2.4) | 15 (5.6) | 0.124 | 2 (2.5) | 6 (2.5) | 0.099 | 3 (3.8) | 17 (7.1) | 0.283 |
| <60 years | 4 (2.6) | 15 (6.1) | 0.113 | 2 (2.8) | 6 (2.8) | 0.099 | 3 (4.2) | 17 (7.9) | 0.277 |
| ≥60 years | 0 (0.0) | 0 (0.0) | --- | 0 (0.0) | 0 (0.0) | --- | 0 (0.0) | 0 (0.0) | --- |
| Erythema (%) | 3 (1.8) | 10 (3.7) | 0.386 | 2 (2.5) | 3 (1.3) | 0.602 | 2 (2.5) | 10 (4.2) | 0.737 |
| <60 years | 3 (2.0) | 10 (4.1) | 0.385 | 1 (1.4) | 3 (1.4) | 0.999 | 1 (1.4) | 10 (4.7) | 0.301 |
| ≥60 years | 0 (0.0) | 0 (0.0) | --- | 1 (12.5) | 0 (0.0) | 0.242 | 1 (12.5) | 0 (0.0) | 0.242 |
| Swelling (%) | 3 (1.8) | 5 (1.9) | 0.999 | 1 (1.3) | 5 (2.1) | 0.999 | 1 (1.3) | 9 (3.8) | 0.461 |
| <60 years | 3 (2.0) | 5 (2.0) | 0.999 | 1 (1.4) | 4 (1.9) | 0.999 | 1 (1.4) | 8 (3.7) | 0.458 |
| ≥60 years | 0 (0.0) | 0 (0.0) | --- | 0 (0.0) | 1 (4.0) | 0.999 | 0 (0.0) | 1 (4.0) | 0.999 |
| **Systemic reactions** |  |  |  |  |  |  |  |  |  |
| Headache (%) | 50 (30.5) | 107 (39.6) | 0.055 | 15 (18.8) | 46 (19.2) | 0.922 | 39 (48.8) | 116 (48.5) | 0.974 |
| <60 years | 49 (32.2) | 102 (41.6) | 0.061 | 12 (16.7) | 42 (19.6) | 0.579 | 36 (50.0) | 109 (50.9) | 0.891 |
| ≥60 years | 1 (8.3) | 5 (20.0) | 0.641 | 3 (37.5) | 4 (16.0) | 0.32 | 3 (37.5) | 7 (28.0) | 0.673 |
| Fatigue (%) | 32 (19.5) | 58 (21.5) | 0.624 | 10 (12.5) | 25 (10.5) | 0.613 | 22 (27.5) | 64 (26.8) | 0.9 |
| <60 years | 31 (20.4) | 55 (22.4) | 0.629 | 8 (11.1) | 23 (10.7) | 0.932 | 20 (27.8) | 60 (28.0) | 0.966 |
| ≥60 years | 1 (8.3) | 3 (12.0) | 0.999 | 2 (25.0) | 2 (8.0) | 0.241 | 2 (25.0) | 4 (16.0) | 0.616 |
| Myalgia (%) | 23 (14.0) | 48 (17.8) | 0.305 | 9 (11.3) | 19 (7.9) | 0.367 | 19 (23.8) | 54 (22.6) | 0.831 |
| <60 years | 22 (14.5) | 46 (18.8) | 0.269 | 8 (11.1) | 16 (7.5) | 0.336 | 18 (25.0) | 50 (23.4) | 0.778 |
| ≥60 years | 1 (8.3) | 2 (8.0) | 0.999 | 1 (12.5) | 3 (12.0) | 0.999 | 1 (12.5) | 4 (16.0) | 0.999 |
| Diarrhea (%) | 18 (11.0) | 36 (13.3) | 0.471 | 5 (6.3) | 18 (7.5) | 0.701 | 15 (18.8) | 44 (18.4) | 0.946 |
| <60 years | 17 (11.2) | 36 (14.7) | 0.318 | 4 (5.6) | 16 (7.5) | 0.58 | 14 (19.4) | 42 (19.6) | 0.973 |
| ≥60 years | 1 (8.3) | 0 (0.0) | 0.324 | 1 (12.5) | 2 (8.0) | 0.999 | 1 (12.5) | 2 (8.0) | 0.999 |
| Nausea (%) | 18 (11.0) | 25 (9.3) | 0.562 | 3 (3.8) | 9 (3.8) | 0.999 | 11 (13.8) | 27 (11.3) | 0.558 |
| <60 years | 18 (11.8) | 22 (9.0) | 0.357 | 2 (2.8) | 9 (4.2) | 0.736 | 10 (13.9) | 24 (11.2) | 0.544 |
| ≥60 years | 0 (0.0) | 3 (12.0) | 0.537 | 1 (12.5) | 0 (0.0) | 0.242 | 1 (12.5) | 3 (12.0) | 0.999 |
| Arthralgia (%) | 10 (6.1) | 14 (5.2) | 0.687 | 2 (2.5) | 7 (2.9) | 0.999 | 7 (8.8) | 18 (7.5) | 0.726 |
| <60 years | 10 (6.6) | 13 (5.3) | 0.596 | 2 (2.8) | 6 (2.8) | 0.999 | 7 (9.7) | 16 (7.5) | 0.544 |
| ≥60 years | 0 (0.0) | 1 (4.0) | 0.999 | 0 (0.0) | 1 (4.0) | 0.999 | 0 (0.0) | 2 (8.0) | 0.999 |
| Anorexia (%) | 10 (6.1) | 18 (6.7) | 0.815 | 3 (3.8) | 3 (1.3) | 0.169 | 6 (7.5) | 16 (6.7) | 0.806 |
| <60 years | 10 (6.6) | 16 (6.5) | 0.985 | 2 (2.8) | 3 (1.4) | 0.603 | 5 (6.9) | 14 (6.5) | 0.999 |
| ≥60 years | 0 (0.0) | 2 (8.0) | 0.999 | 1 (12.5) | 0 (0.0) | 0.242 | 1 (12.5) | 2 (8.0) | 0.999 |
| Pruritus (%) | 2 (1.2) | 10 (3.7) | 0.225 | 0 (0.0) | 4 (1.7) | 0.575 | 1 (1.3) | 14 (5.9) | 0.127 |
| <60 years | 2 (1.3) | 9 (3.7) | 0.217 | 0 (0.0) | 4 (1.9) | 0.575 | 1 (1.4) | 13 (6.1) | 0.202 |
| ≥60 years | 0 (0.0) | 1 (4.0) | 0.999 | 0 (0.0) | 0 (0.0) | --- | 0 (0.0) | 1 (4.0) | 0.999 |
| Exanthema (%) | 1 (0.6) | 7 (2.6) | 0.268 | 0 (0.0) | 1 (0.4) | 0.999 | 1 (1.3) | 8 (3.3) | 0.459 |
| <60 years | 1 (0.7) | 7 (2.9) | 0.161 | 0 (0.0) | 1 (0.5) | 0.999 | 1 (1.4) | 8 (3.7) | 0.458 |
| ≥60 years | 0 (0.0) | 0 (0.0) | --- | 0 (0.0) | 0 (0.0) | --- | 0 (0.0) | 0 (0.0) | --- |
| Allergy (%) | 1 (0.6) | 6 (2.2) | 0.262 | 0 (0.0) | 3 (1.3) | 0.575 | 0 (0.0) | 8 (3.3) | 0.209 |
| <60 years | 0 (0.0) | 5 (2.0) | 0.161 | 0 (0.0) | 2 (0.9) | 0.999 | 0 (0.0) | 4 (1.9) | 0.575 |
| ≥60 years | 1 (8.3) | 1 (4.0) | 0.999 | 0 (0.0) | 1 (4.0) | 0.999 | 0 (0.0) | 1 (4.0) | 0.999 |
| Vomiting (%) | 3 (1.8) | 1 (0.4) | 0.154 | 0 (0.0) | 4 (1.7) | 0.575 | 0 (0.0) | 4 (1.7) | 0.575 |
| <60 years | 3 (2.0) | 1 (0.4) | 0.159 | 0 (0.0) | 4 (1.9) | 0.575 | 0 (0.0) | 4 (1.9) | 0.575 |
| ≥60 years | 0 (0.0) | 0 (0.0) | --- | 0 (0.0) | 0 (0.0) | --- | 0 (0.0) | 0 (0.0) | --- |
| Fever (>37.8°C) (%) | 1 (0.6) | 1 (0.4) | 0.999 | 0 (0.0) | 0 (0.0) | --- | 1 (1.3) | 1 (0.4) | 0.439 |
| <60 years | 1 (0.7) | 1 (0.4) | 0.999 | 0 (0.0) | 0 (0.0) | --- | 1 (1.4) | 1 (0.5) | 0.441 |
| ≥60 years | 0 (0.0) | 0 (0.0) | --- | 0 (0.0) | 0 (0.0) | --- | 0 (0.0) | 0 (0.0) | --- |

Percentages were calculated from the total number of participants in each group.
Data in the table were reported within 7 days post any of the two doses.
Sample sizes: Placebo <60 1<sup>st</sup> dose: n=152; Placebo <60 2<sup>nd</sup> dose: n=72; Vaccine <60 1<sup>st</sup> dose: n=245; Vaccine <60 2<sup>nd</sup> dose: n=214; Placebo ≥ 60 1<sup>st</sup> dose: n=12 ; Placebo ≥60 2<sup>nd</sup> dose: n=8 ; Vaccine ≥60 1<sup>st</sup> dose: n=25; Vaccine ≥60 years 2<sup>nd</sup> dose: n=25.

A total of 55 unsolicited AEs were reported. Those with a frequency of 1% or more were gastrointestinal discomfort (n=7), abdominal pain (n=5), odynophagia (n=5), and back pain (n=4). During the study period, three COVID-19 cases occurred (breakthrough cases). Two were detected at enrolment: one of them developed anosmia in the following days, and another after the first dose. One of them had a clinical progression score of 1 (asymptomatic), and the other two had a score of 2 (symptomatic, independent) ^14^. No SAEs nor other events of special interest occurred in this study period.

### 2. Immunization with CoronaVac in a 0-14 schedule induces the secretion of IgG against the S1-RBD of SARS-CoV-2 with potentially neutralizing capacities in adults aged 18-59 and ≥60

Evaluation of IgG specific against S1-RBD and the N protein of SARS-CoV-2 was performed independently using ELISA assays (Fig. S1 and S2). A total of 32 participants aged 18-59 (23 in the vaccine arm and 9 in the placebo arm) and 14 participants aged ≥60 were evaluated (11 in the vaccine arm and 3 in the placebo arm). Four of the participants aged 18-59 were identified as seropositive at the time of recruitment. Seroconversion rates for the S1-RBD specific IgG (Table 3 - Upper) were 47.8% 14 days p.i. in participants aged 18-59 (GMT 23.1) and 18.1% at the same time point in participants aged ≥60 (GMT 45.1). At 28 and 42 days p.i., seroconversion rates reached 95.6% in participants aged 18-59 (GMT 1,755 and 1,878 at day 28 and 42 p.i., respectively). Seroconversion rates in participants aged ≥60 reached 100% at day 28 p.i. (GMT 1,860.2) and 87.5% at day 42 p.i. (GMT 1,878) (Table 3 and Figure 2). Interestingly, the seroconversion rates for IgG specific against the N protein (Table 3 – Middle) 14 days p.i. was 8.7% for participants aged 18-59 (GMT 5.3) and 0% for participants aged ≥60 (GMT 5.5). On day 28 p.i., the seroconversion rates were 17.4% for the 18-59 years old group and 20.0% ≥60 years old group (GMT 8.0 and 9.6 respectively), which increased up to 37.5% for the participants ≥60 years old (GMT 32.6) and decreased to 13.0% in participants aged 18-59 (GMT 9.2) 42 days p.i. (Table 3 and Figure 2). The results obtained for the seropositive participants at enrollment and breakthrough cases are shown in Table S5. These results suggest that CoronaVac induces a significant production of S1-RBD specific IgG after immunization with a 0-14 scheme but induces a weak production of IgG specific against the N protein. We confirmed that doses of CoronaVac contain significant amounts of the N protein (Fig. S3).

**Table 3.**
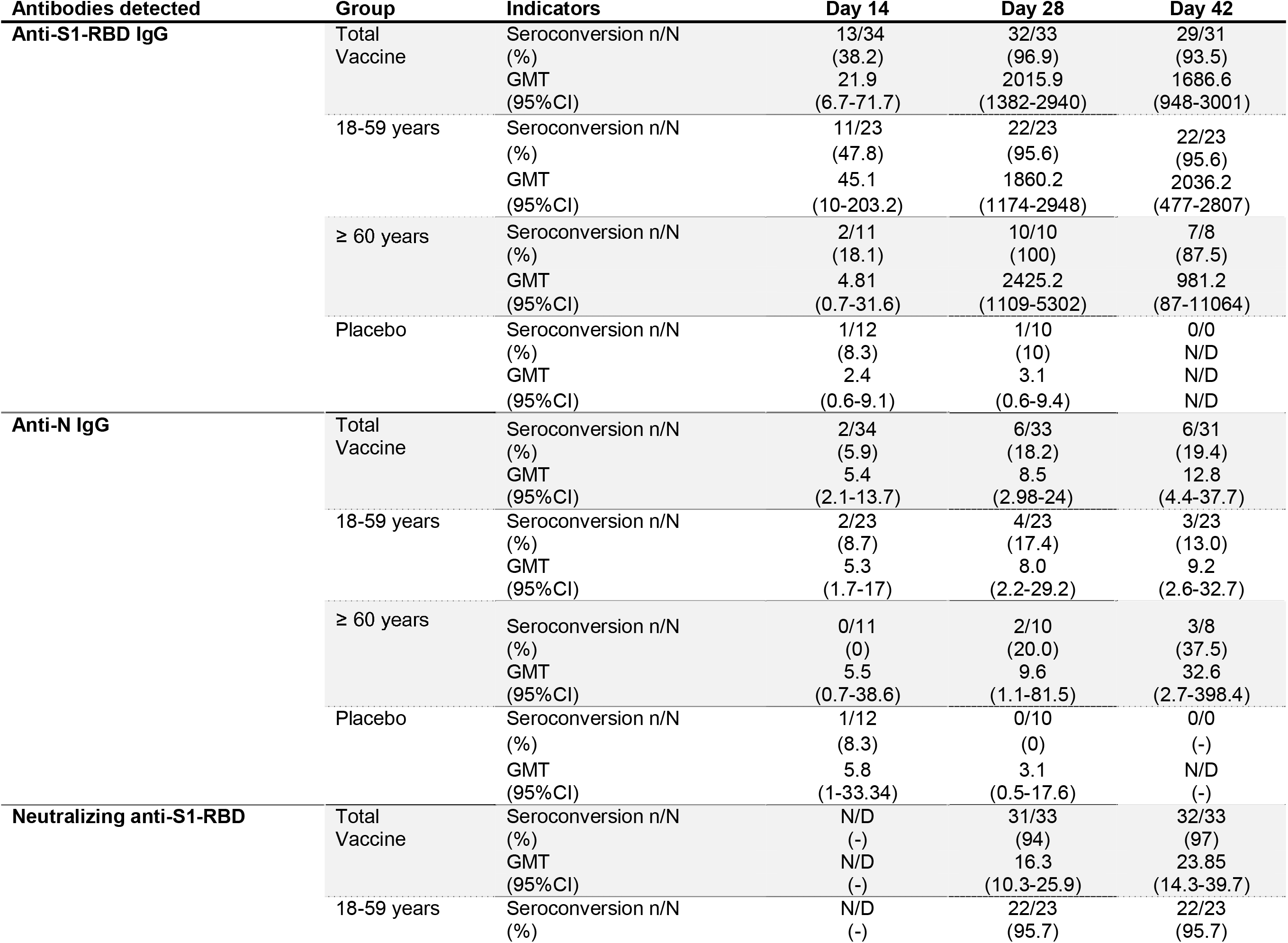
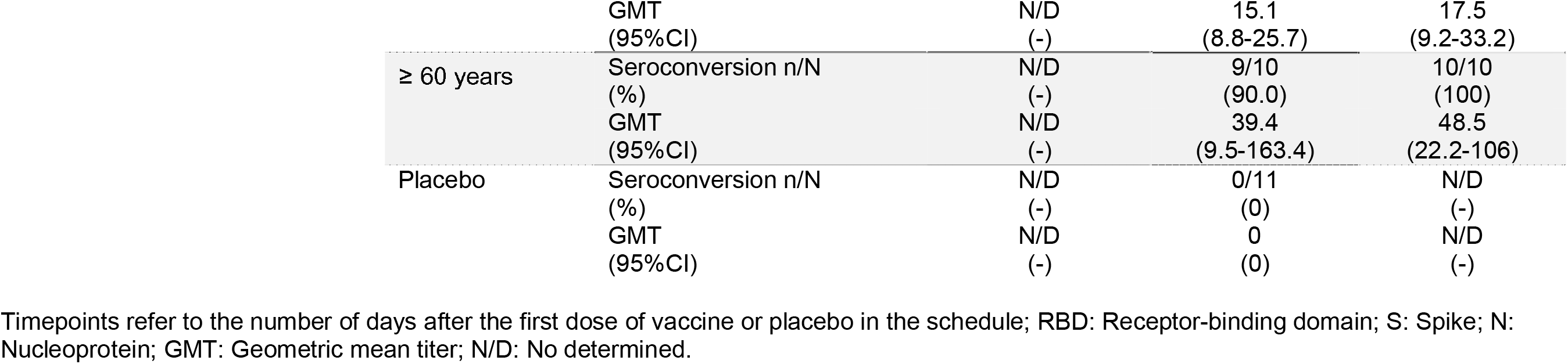
**Seroconversion rates and Geometric median titer of antibodies against SARS-CoV-2 proteins**

**Figure 2:**
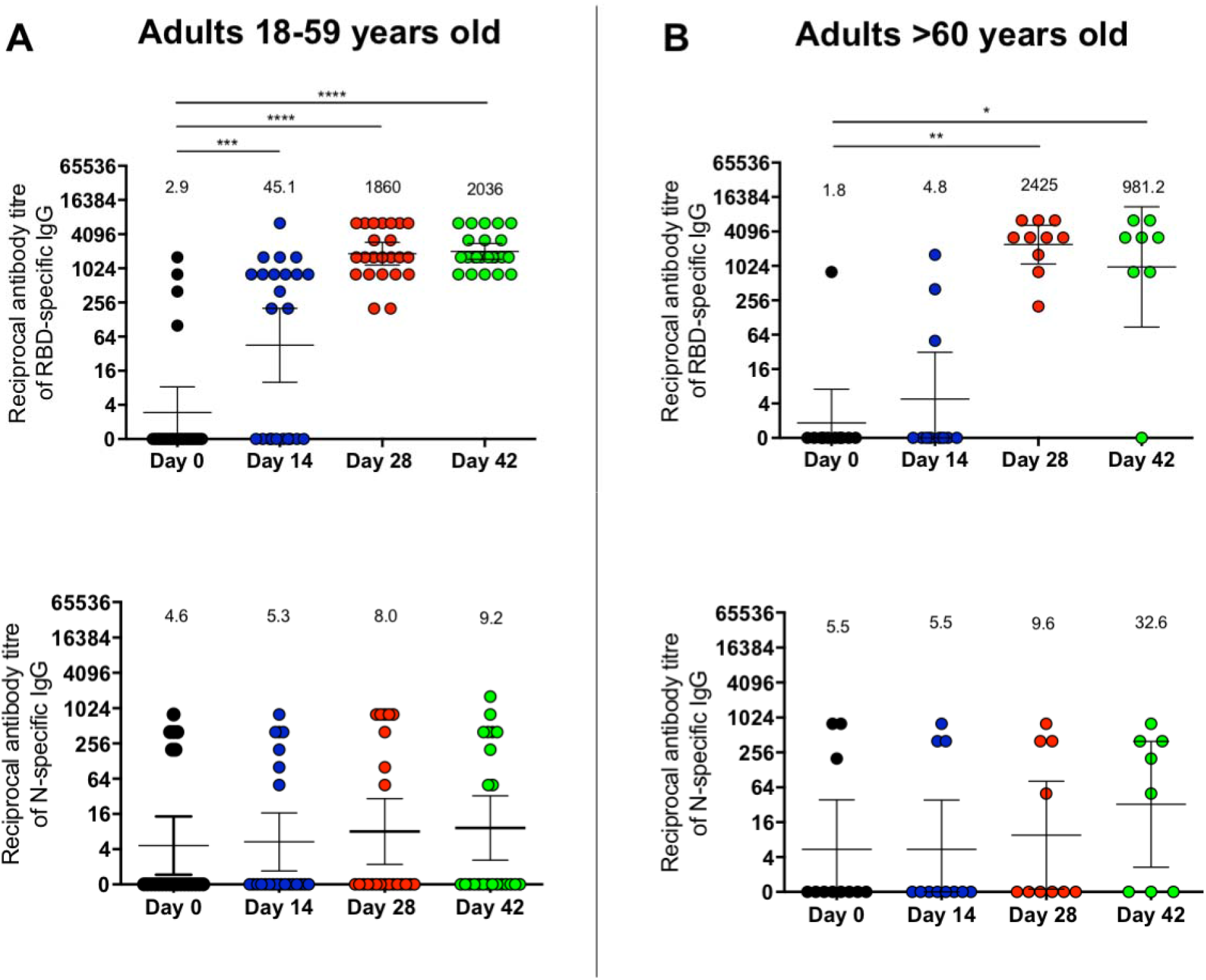
Immunization with CoronaVac induces specific IgG against SARS-CoV-2 antigens in participants aged 18-59 and ≥60 after two immunizations in a 0-14 schedule. Titers of IgG antibodies after two doses of CoronaVac were evaluated for immunized participants (excluding seropositive participants at recruitment and placebo participants) at 0, 14, 28, and 42 days post the first immunization (p.i.) for adults aged 18-59 (**A, C**), and ≥60 (**B, D**) for specific IgG against the S1-RBD (upper panel) and the N protein (lower panel) of SARS-CoV-2. Data are expressed as the reciprocal antibody titer v/s time after the first dose. Error bars indicate the 95% CI of the Geometric Mean Titer (GMT). The spots represent the individual values of antibody titers, with the numbers above the spots showing the GMT estimates. The graph illustrates the results obtained for 23 participants in the 18-59 years old group and 11 participants in the ≥60 years old group. A Wilcoxon test was performed to compare the samples of day 0 against the rest of the groups; *p<0.05, **p<0.005, ***p<0.0005, ****p<0.0001.

To evaluate the potential neutralizing capacities of these antibodies, a sVNT was performed. This test detects whether antibodies present in the sera interfere with the binding of S1-RBD to the hACE2 receptor. Serum samples used were obtained from 44 participants aged 18-59 (23 in the vaccine arm and 8 in the placebo arm) and 14 participants aged ≥60 (10 in the vaccine arm and 3 in the placebo arm), at 0, 28, and 42 days p.i.. An inhibition rate of ≤30% was set as the negative cut-off for this assay. We observed that the titers of antibodies against the RBD region increased significantly 28 and 42 days p.i. as compared to day 0. The GMTs calculated for the antibody titers in participants aged 18-59 were 15.1 on day 28 p.i., and 17.5 on day 42 p.i., (Fig. 3A). Regarding the ≥60 years old group, the GMTs were 39.4 and 48.5, at 28 and 42 days p.i., respectively (Fig. 3B). The seroconversion rate was 96% for the 18-59 age group at 28 and 42 days p.i. and 90% and 100% for the ≥60 age group at these times. The neutralizing capacities of antibodies induced by vaccination were also evaluated by three additional methodologies (pseudotyped virus assay^15^, BioHermes surrogate VNT and neutralization of live SARS-CoV-2), showing similar results (Fig. S4). These results suggest that immunization with CoronaVac in a 0-14 schedule promotes the production of anti-S1-RBD IgG with neutralizing capacities in both age groups.

**Figure 3.**
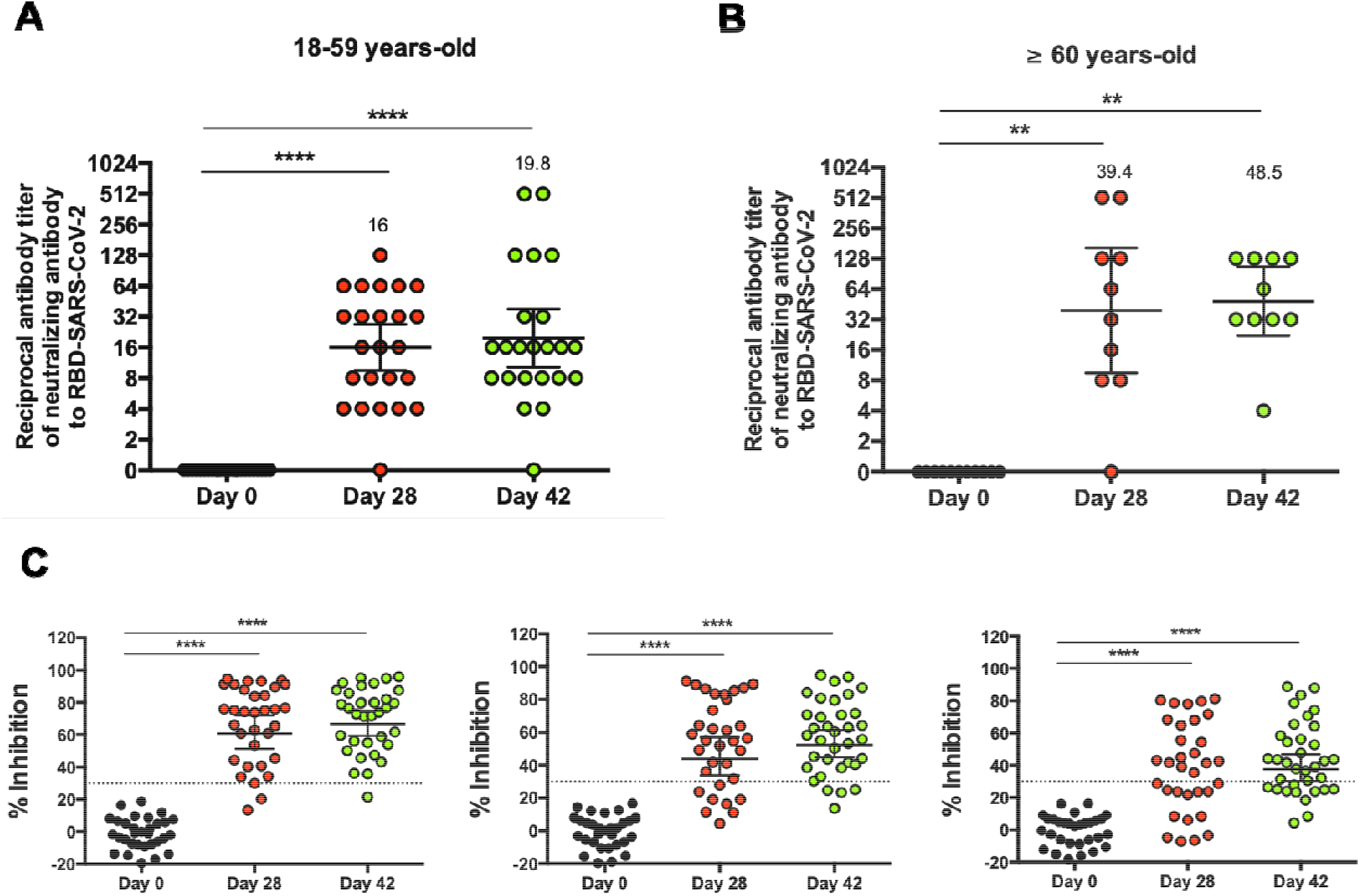
Immunization with CoronaVac induces neutralizing antibodies against SARS-CoV-2 in participants aged 18-59 and ≥60 after two immunizations in a 0-14 schedule. Neutralizing antibody titers were evaluated with a surrogate virus neutralization assay, which quantifies the interaction between S1-RBD and hACE2 pre-coated on ELISA plates. Results were obtained from participants aged (A) 18-59 and (B) ≥ 60 at 0, 28, and 42 days p.i. Data is represented as the reciprocal antibody titer v/s time after the first dose. Numbers above the bars show the Geometric Mean Titer (GMT), and the error bars indicate the 95% CI. Data were analyzed by a Wilcoxon test to compare against day 0; *p<0.05, **p<0.005, ***p<0.0005, ****p<0.0001. (C) Percentage of inhibition of the neutralizing antibodies at 1:4, 1:8, and 1:16 dilutions of sera from vaccinated participants. The graph represents the results obtained for 23 participants in the 18-59 years old group and 10 participants in the ≥60 years old group. Dotted lines indicate the cut-off value, set at 30%. Data were analyzed by a two-tailed Student’s t-test against day 0; ****p< 0.0001

### 3. Immunization with CoronaVac in a 0-14 schedule induces IFN-γ-producing T cells specific for SARS-CoV-2 antigens in adults aged 18-59 and ≥60 in the Chilean population

To evaluate the cellular immune response elicited upon vaccination with CoronaVac, the specific T cell responses against MP of peptides derived from the S protein of SARS-CoV-2 (MP-S) and the remaining proteins of this virus (MP-R) were evaluated by ELISPOT and flow cytometry assays in a total of 36 participants from the vaccine arm (27 aged 18-59 and 9 aged ≥60) and 6 from the placebo arm. We observed an increase in the number of Spot Forming Cells (SFCs) for IFN-γ at 28 and 42 days p.i. as compared to day 0, upon stimulation with both the MP-S and the MP-R (Fig. 4A and B). This tendency was seen for participants aged 18-59 and ≥60 (Fig. 4). Similar trends were observed when a fold change analysis of this increase was performed and normalized to day 0 (Fig. S5). In the 18-59 age group, the number of SFCs for IFN-γ producing T cells showed an average increase of 14.04 and 9.76 times for the MP-S and 31.78 and 18.67 times for the MP-R at 28 and 42 days p.i., respectively. In the ≥60 years old group, an average increase of 20.04 and 9.63 times was observed for the MP-S, and 33.81 and 20.28 times for the MP-R at 28 and 42 days p.i., respectively. Participants in the placebo arm exhibited no differences in the number of SFCs for IFN-γ among the different stimuli and time points evaluated (Fig. S6). The response detected in the ELISPOT assays suggests that immunization with CoronaVac induces a T cell response polarized towards a Th1 immune profile, as the secretion of IL-4 by T cells was mainly undetected (Fig. S7). As a positive control, PBMCs from participants were stimulated with an MP of peptides derived from cytomegalovirus (CMV) (Fig. S8).

**Figure 4.**
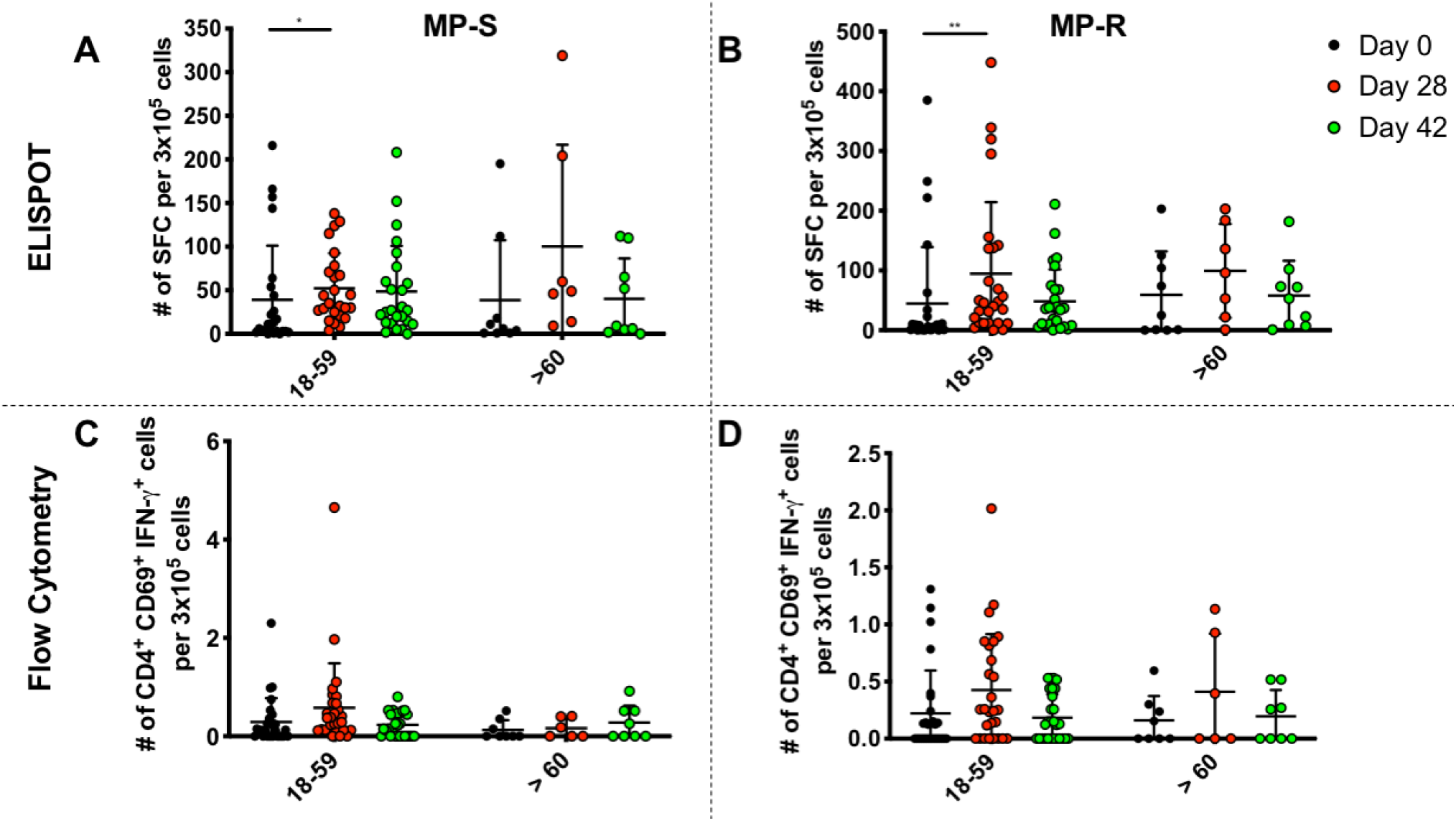
Quantification of IFN-γ-secreting CD4^+^ T cells, upon stimulation with two Mega Pools of peptides derived from SARS-CoV-2 proteins (MP-S and MP-R) in participants aged 18-59 and ≥60 immunized with CoronaVac. Absolute count of IFN-γ**-**secreting cells, determined by ELISPOT as Spot Forming Cells (SFCs), was measured upon stimulation of PBMC with MP-S (A) and MP-R (B), for 48 h in samples obtained at 0, 28, and 42 days p.i. The absolute number of activated CD4^+^ T cells (defined as CD3^+^, CD69^+^) secreting IFN-γ, as determined by flow cytometry, was measured upon stimulation for 24 h with MP-S (C) and MP-R (D) in samples obtained at 0, 28, and 42 days p.i. A total of 25 samples stimulated with MP-S and 27 samples stimulated with MP-R were considered in the 18-59 years old group and 9 samples for the ≥60 years old group. Data shown represent means ± SD. Data from each age group were analyzed separately by a Friedman test for repeated measures, followed by a *post hoc* Dunn’s test corrected for multiple comparisons against day 0 for each age group; *=p<0.05, **=p<0.005.

Since MP-S and MP-R were originally determined *in silico* to optimally stimulate CD4^+^ T cells, the activation and the secretion of Th1 cytokines by these cells upon stimulation were evaluated by flow cytometry. Remarkably, the absolute numbers of activated CD4^+^ T cells (defined as CD3^+^, CD4^+^, CD69^+^) increased at 28 days p.i. as compared to day 0 in both age groups (Fig. S9A and Table S6). The absolute number of the activated CD4^+^ T cells secreting IFN-γ showed an evident increase at 28 days p.i. in the 18-59 years old group, upon stimulation with both MP-S and MP-R (Fig. 4C and D). However, this trend was not evident for the ≥60 years old group, primarily upon stimulation with the MP-S. A fold change analysis of the activated CD4^+^ T cells secreting IFN-γ at days 28 and 42 p.i., compared to day 0, also suggests that an increase is observed for the 18-59 years old group, but not for the ≥60 years old group (Table S6). We also evaluated the absolute numbers (Fig. S10) and the fold change (Table S6) of activated CD4^+^ T cells producing IL-2 or TNF-α upon stimulation with MP-S and MP-R, evidencing variable trends among both age groups, 28 and 42 days p.i., compared to day 0.

The specific T cell responses against MP of peptides chosen from the whole proteome of SARS-CoV-2 (MP-CD8A and MP-CD8B) was also evaluated in 7 participants aged 18-59 and 7 participants aged ≥60. In most of the participants evaluated for the 18-59 age group, stimulation with MP-CD8A and CD8B resulted in a modest increase in SFCs for IFN-γ 28 and 42 days p.i. compared to day 0 (Fig. 5A and B). However, a partial rise in SFCs producing IFN-γ was observed upon stimulation with both MP-CD8A and MP-CD8B for some participants aged ≥60 (Fig. 5B), at 28 and 42 days p.i.. There was a subtle fold increase in the number of SFCs positive for IFN-γ in participants aged 18-59 and a more evident fold increase in participants aged ≥60 with both MP, at 28 and 42 days p.i. (Fig. S11). No marked differences were detected for the placebo arm participants among the different stimuli and times evaluated (Fig. S12).

**Figure 5.**
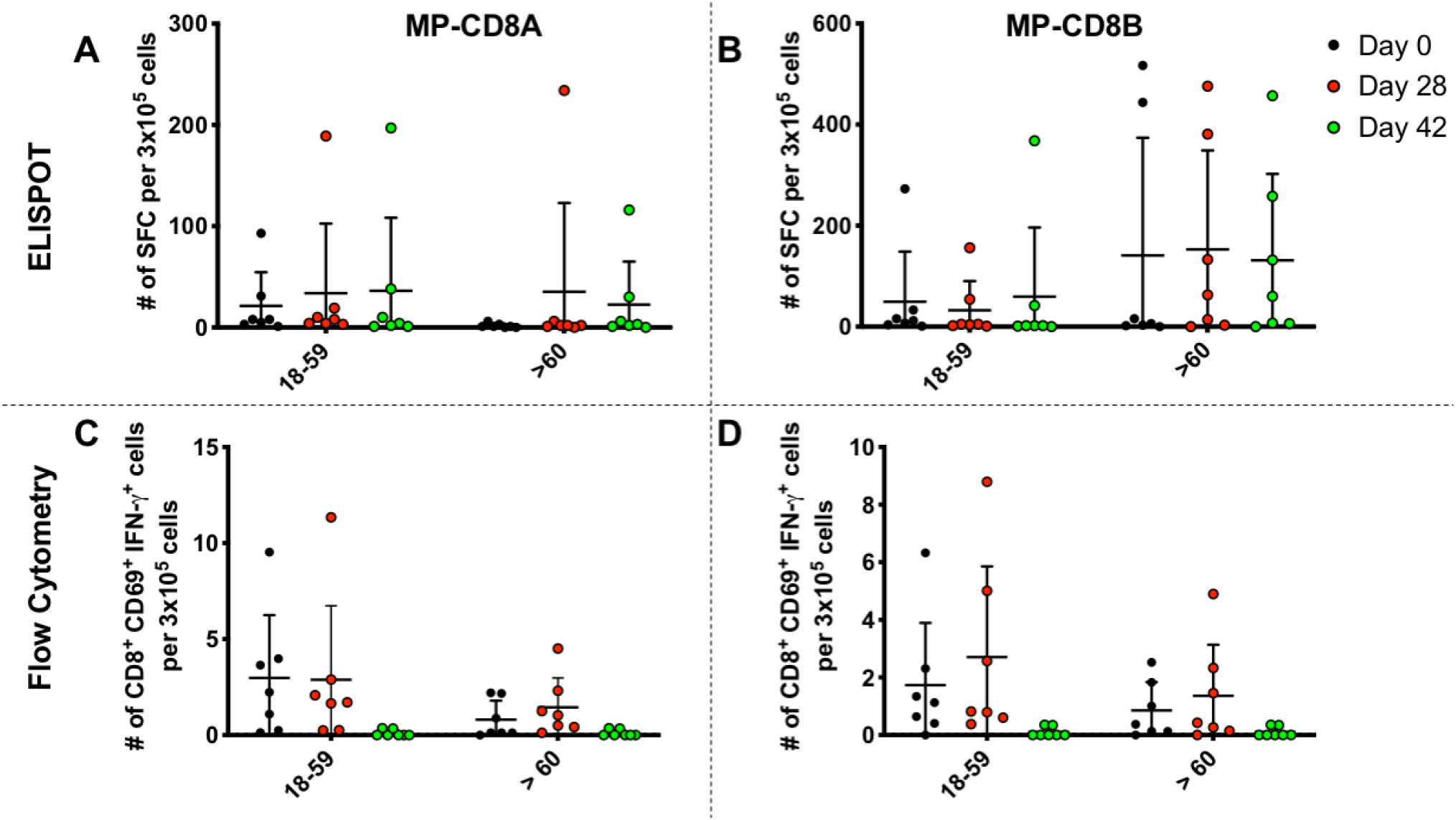
Quantification of IFN-γ-secreting CD8^+^ T cells, upon stimulation with two Mega Pools of peptides derived from SARS-CoV-2 proteins (MP-CD8A and CD8B) in participants aged 18-59 and ≥60, immunized with CoronaVac. Absolute count of IFN-γ**-** secreting cells, determined by ELISPOT as Spot Forming Cells (SFCs), was measured upon stimulation of PBMC with MP-CD8A (**A**) and MP-CD8B (**B**) for 48 h in samples obtained at 0, 28, and 42 days p.i.. The absolute number of activated CD8^+^ T cells (defined as CD3^+^, CD69^+^) secreting IFN-γ, determined by flow cytometry, was measured upon stimulation for 24 h with MP-CD8A (**C**) and MP-CD8B (**D**) in samples obtained at 0, 28, and 42 days p.i.. A total of 7 participants were considered in the 18-59 years old group and 7 participants for the ≥60 years old group. Data shown represent mean ± SD. Data from each age group were analyzed separately by a Friedman test for repeated measures, followed by a *post hoc* Dunn’s test corrected for multiple comparisons against day 0 for each age group.

Since the peptides included in MP-CD8A and MP-CD8B were determined *in silico* to stimulate CD8^+^ T cells, the activation and the secretion of Th1 cytokines by these cells upon stimulation was measured by flow cytometry, as described in ^13^. Variable changes were seen in the activation of CD8^+^ T cells (defined as CD3^+^, CD8^+^, CD69^+^), and the secretion of cytokines by these cells. For both groups, the absolute number of activated CD8^+^ T cells showed no significant variation at 28 and 42 days p.i. compared to day 0 (Fig. S13). Minor increases at 28 days p.i. were observed for the absolute numbers of CD8^+^ T cells that produced the cytokines IFN-γ in both age groups (Fig. 5C and D). Fold change analys of these results can be found on Table S6. We also evaluated absolute numbers (Fig. S14) and fold increases (Table S6) of activated CD8^+^ T cells producing IL-2 or TNF-α upon stimulation with MP-CD8A and MP-CD8B. Variable trends for these cells at 28 and 42 days p.i. for both groups were detected, as compared to day 0. Although a higher number of participants must be evaluated, ELISPOT and flow cytometry results suggest that stimulation with MP-CD8A and MP-CD8B can, to some extent, induce a cellular immune response in participants immunized with CoronaVac.

Overall, these results suggest that CoronaVac can induce a humoral immune response based on total and neutralizing antibodies and a CD4^+^ T cell response polarized towards a Th1 profile in adults aged 18 and older, which targets several SARS-CoV-2 antigens.

## Discussion

This study is a first preliminary analysis of a phase 3 clinical trial performed in Chile with CoronaVac, an inactivated vaccine against SARS-CoV-2. Results show that this vaccine has a favorable safety and immunogenicity profile in the Chilean population. Although further analyses including more participants are needed, the results shown here are similar to those obtained in previous clinical trials with CoronaVac ^10,11^. We found that two doses of CoronaVac, in a 0-14 schedule, were safe and capable of inducing a humoral and cellular immune response in both age groups evaluated (18-59 and ≥60 years old). A different immunization schedule considering a booster at day 28 p.i. instead of day 14 p.i. is being currently tested, in line with results recently reported on the Chinese population^10^. Further studies will focus on this new immunization schedule.

Adverse reactions observed were primarily mild and local, which coincides with previous reports with this vaccine. The most reported solicited local AE was pain at the injection site in the vaccine arm resolved within two days after vaccination. Headache was the most systemic common solicited AE registered in the vaccine and the placebo arm, at a similar frequency. No SAEs were reported for either the vaccine or placebo arm. Interestingly, we detected differences between the age groups regarding the frequency of local and systemic AEs. These were more frequent in the 18-59 years old group than in the ≥60 years old group.

The antibody response elicited by the Chilean population was similar to the one reported previously. We detected over 90% seroconversion for S1-RBD-specific IgG, along with over 90% of seroconversion for neutralizing antibodies at 28 and 42 days p.i.. These rates are consistent with the data reported in the phase 2 trial conducted in China (100% for RBD-specific IgG and 99.2% for neutralizing antibodies) for the same immunization scheme, dose, and age ^10^. Although seroconversion rates are slightly lower in participants aged ≥60 at 42 days p.i., these results are promising, as the vaccine may exhibit a protective profile in older populations. Differences in the seroconversion rates and antibodies neutralizing capacities seen among the different methodologies tested may be directly related with the intrinsic characteristics of each methodology (i.e. sVNA, pseudotype virus neutralization assays, and infection inhibition).

We observed a low secretion of anti-N antibodies as compared to IgG induced against the S1-RBD, which is not related to the absence of the N protein in CoronaVac. Previous reports indicate that humans naturally infected with SARS-CoV-2 develop antibody responses mainly to the S and N proteins, in somewhat similar levels ^9^. However, immunization studies of mice, rats, and non-human primates with CoronaVac showed that the antibodies induced were mostly directed against the S protein and the S1-RBD, with a reduced amount of antibodies against the N protein ^9^. This is in line with our findings, suggesting that the enhanced secretion of antibodies against the S protein by CoronaVac, rather than against the N protein, may be playing a role in the protective response induced by the vaccine. This may also have significant considerations when choosing techniques for the confirmation of infections with SARS-CoV-2, as anti-N antibodies may not be mostly detected in immunized individuals.

This is the first time a characterization of the cellular response against proteins other than the S protein of SARS-CoV-2 is reported in humans immunized with CoronaVac. Unlike previous studies^10^, we detected a robust T cell response upon stimulation of PBMCs with MPs of peptides from S (MP-S) and the remaining viral particle (MP-R). There was a clear increase in the number of SFCs for IFN-γ when PBMC from days 28 and 42 p.i. were stimulated with MP-S and MP-R, which was also evident in the fold changes calculated compared to day 0. An increase in the number of activated CD4^+^ T cells secreting IFN-γ upon stimulation with these stimuli in those time points was detected. However, a reduced number was observed in the ≥60 years old group stimulated with MP-S. This observation could be explained by a natural reduction of activated CD4^+^ T cells in the older population, previously described for other vaccines ^16^. We also evaluated the response elicited upon stimulation with two MPs of peptides designed to stimulate a CD8^+^ T cell response (MP-CD8A and MP-CD8B). Stimulation with MP-CD8 (A and B) resulted in increased SFCs for IFN-γ in some participants in the 18-59 years old group and the ≥60 years old group. We did not observe an evident increase in the number of activated CD8^+^ T cells secreting IFN-γ at days 28 and 42 p.i. in none of the age groups. Although more participants will be evaluated to raise more robust conclusions, the results obtained so far suggest that the CD8^+^ immune response detected in vaccinated participants is not as robust as the CD4^+^ immune response. Since the secretion of IFN-γ by CD4^+^ T cells and reduced amounts of IL-4 secreting cells are in line with a well-balanced immune response that could achieve virus clearance, immunization with CoronaVac shows promising capacities of inducing an antiviral response in the host. This IFN-γ response has also been sought and observed in other vaccines against SARS-CoV-2, such as BNT162b1 designed by BionTech ^17^ and a recombinant adenovirus type-5 vectored COVID-19 vaccine designed by CanSino ^18^.

In summary, immunization with CoronaVac is safe and induces robust humoral and cellular responses, characterized by increased antibody titers against the S1-RBD with neutralizing capacities, and the production of T cells that are specific for several SARS-CoV-2 antigens and were characterized by the secretion of Th1 cytokines.

## Supporting information

Supplementary Annex

## Data Availability

All the data included in the manuscript is available upon request to the corresponding authors.

## Acknowledgments

This study was supported by the Ministry of Health, Government of Chile; The Confederation of Production and Commerce, Chile; the Consortium of Universities for Vaccines and Therapies against COVID-19, Chile; the Chilean Public Health Institute (ISP); and the Millennium Institute on Immunology and Immunotherapy. NIH contracts 75N93019C00001(A.S) and 75N9301900065 supported A.S, D.W. We would like to thank the support of the Ministry of Science, Technology, Knowledge, and Innovation, Government of Chile; and The Ministry of Foreign Affairs, Government of Chile. We also would like to thank PATH for sharing the First WHO International Standard for anti-SARS-CoV-2 immunoglobulin;Alex Cabrera and Sergio Bustos from the Flow Cytometry Facility at Facultad de Ciencias Biológicas, Pontificia Universidad Católica de Chile for support with flow cytometry; and Sebastián Silva from the same Faculty for support with statistical analyses. We also thank the Vice Presidency of Research (VRI), the Direction of Technology Transfer and Development (DTD), the Legal Affairs Department (DAJ) and the Administrative Direction from the School of Biological Sciences and the School of Medicine of the Pontificia Universidad Católica de Chile. Special thanks to the members of the CoronaVac03CL Study Team (list in Supplementary Appendix) for their hard work and dedication to the clinical study; the members of the independent data safety monitoring committee (members in Supplementary Appendix) for their oversight, and the subjects enrolled in the study for their participation and commitment to this trial.

