## Supplementary material for "Interim report: Safety and immunogenicity of an inactivated vaccine against SARS-CoV-2 in healthy chilean adults in a phase 3 clinical trial"

**Brief Title:** CoronaVac03CL Phase 3 Interim Analysis of Safety and Immunogenicity.

Susan M Bueno^1,2^, Katia Abarca^1,3^, Pablo A González^1,2^, Nicolás MS Gálvez^1,2^, Jorge A Soto^1,2^, Luisa F Duarte^1,2^, Bárbara M Schultz^1,2^, Gaspar A Pacheco^1,2^, Liliana A González^1,2^, Yaneisi Vázquez^1,2^, Mariana Ríos^1,2^, Felipe Melo-González^1,2^, Daniela Rivera^1,2^, Carolina Iturriaga^3^, Marcela Urzúa^3^, Angélica Dominguez^4^, Catalina A Andrade^1,2^, Roslye V Berrios^1,2^, Gisela Canedo-Marroquín^1,2^, Camila Covián^1,2^, Farides Saavedra^1,2^, Omar P Vallejos^1,2^, Paulina Donato^5^, Pilar Espinoza^6,7^, Daniela Fuentes^8^, Marcela González^9,10^, Paula Guzmán^11^, Paula Muñoz-Venturelli^12^, Carlos M Pérez^6,7^, Marcela Potin^13^, Alvaro Rojas^14^, , Rodrigo Fasce^15^, Jorge Fernández^15^, Judith Mora^15^, Eugenio Ramírez^15^, Aracelly Gaete-Argel^16^, Aarón Oyarzún-Arrau^16^, Fernando Valiente-Echeverría^16^, Ricardo Soto-Rifo^16^, Daniela Weiskopf^17^, Alessandro Sette^17^, Gang Zheng^18^, Weining Meng^18^, José V González-Aramundiz^19^, Alexis M Kalergis^1,2,20^*

^1^Millennium Institute on Immunology and Immunotherapy.

^2^Departamento de Genética Molecular y Microbiología, Facultad de Ciencias Biológicas, Pontificia Universidad Católica de Chile, Santiago, Chile.

^3^Departamento de Enfermedades Infecciosas e Inmunología Pediátricas, División de Pediatría, Escuela de Medicina, Pontificia Universidad Católica de Chile, Santiago, Chile.

^4^Public Health Department, Facultad de Medicina, Pontificia Universidad Católica de Chile.

^5^Complejo Asistencial Dr. Sótero del Rio, Santiago, Chile.

^6^Hospital Clínico Félix Bulnes.

^7^Facultad de Medicina y Ciencia, Universidad San Sebastián, Santiago, Chile.

^8^Hospital Carlos Van Buren, V Región, Chile.

^9^Hospital Dr. Gustavo Fricke, V Región, Chile.

^10^Departamento de Pediatría, Universidad de Valparaíso.

^11^Clinica Los Andes, Universidad de Los Andes, Santiago, Chile.

^12^Facultad de Medicina, Clínica Alemana Universidad del Desarrollo, Santiago, Chile.

^13^Clinica San Carlos de Apoquindo, Red de Salud UC Christus, Santiago, Chile.

^14^Departamento de Enfermedades Infecciosas del Adulto, División de Medicina, Escuela de Medicina, Pontificia Universidad Católica de Chile, Santiago, Chile.

^15^Departamento de Laboratorio Biomédico, Instituto de Salud Pública de Chile.

^16^Laboratory of Molecular and Cellular Virology, Virology Program, Institute of Biomedical Sciences, Faculty of Medicine, Universidad de Chile, Santiago, Chile.

^17^Center for Infectious Disease and Vaccine Research, La Jolla Institute for Immunology, La Jolla, CA 92037, USA.

^18^Sinovac Biotech, Beijing, China.

^19^Departamento de Farmacia, Facultad de Química y de Farmacia. Pontificia Universidad Católica de Chile, Santiago, Chile.

^20^Departamento de Endocrinología, Facultad de Medicina, Escuela de Medicina, Pontificia Universidad Católica de Chile, Santiago, Chile.

*Corresponding author: Alexis M Kalergis, Pontificia Universidad Católica de Chile. Av. Libertador Bernardo O’Higgins Nº 340, Santiago 8331010, Santiago, Chile. Phone 56-2-6862846, e-mail address:

Keywords: SARS-CoV-2, CoronaVac®, phase 3 clinical trial, safety, immunogenicity, Chile.

| **Table S1. Severity grading criteria for local adverse events** | | | | |
| --- | --- | --- | --- | --- |
| **Adverse Event** | **Grade 1** | **Grade 2** | **Grade 3** | **Grade 4** |
| Pain at the site of the investigational product administration | Does not interfere with daily activities | Repeated use of non-narcotic pain reliever > 24 hours  OR  interferes with activity | Any use of narcotic pain reliever or prevents daily activity | Visit to the emergency room*  OR  Hospitalization |
| Erythema at the site of investigational product administration^†^ | 25 – 50 mm | 51 – 100 mm | > 100 mm | Necrosis  OR  Exfoliative dermatitis |
| Swelling at the site of investigational product administration^†^ | 25 – 50 mm | 51 – 100 mm  OR  interferes with activity | > 100 mm  OR  prevents daily activity | Necrosis |
| Induration at the site of investigational product administration^†^ | 25 – 50 mm | 51 – 100 mm  OR  interferes with activity | > 100 mm  OR  prevents daily activity | Necrosis |
| Pruritus at the site of investigational product administration | Does not interfere with daily activities | Interferes with activity | Prevents daily activity | Visit to the emergency room*  OR  Hospitalization |

| **Table S2. Severity grading criteria for systemic adverse events and vital signs** | | | | | |
| --- | --- | --- | --- | --- | --- |
| **Adverse Event** | | **Grade 1** | **Grade 2** | **Grade 3** | **Grade 4** |
| Fever | 37.8 – 38.4°C | | 38.5 – 38.9°C | 39.0 – 40.0°C | >40°C |
| Nausea | Does not interfere with daily activities  OR  1 to 2 episodes in 24 hours | | Interferes slightly with daily activities  OR  More than 2 episodes in 24 hours | Prevents daily activities, requires intravenous hydration | Visit to the emergency room*  OR  Hospitalization  for hypovolemic shock |
| Vomiting | Does not interfere with daily activities  OR  1 to 2 episodes in 24 hours | | Interferes slightly with daily activities  OR  More than 2 episodes in 24 hours | Prevents daily activities, requires intravenous hydration | Visit to the emergency room*  OR  Hospitalization  OR  Hypovolemic shock |
| Diarrhea | 2 – 3 loose stools in 24 hours | | 4 – 5 stools in 24 hours | 6 or more watery stools or requires outpatient IV hydration | Visit to the emergency room*  OR  Hospitalization |
| Headache | Does not interfere with daily activities | | Repeated use of non-narcotic pain reliever > 24 hours  OR  Interferes slightly with daily activities | Significant; any use of narcotic pain reliever or prevents daily activity | Visit to the emergency room*  OR  Hospitalization |
| Fatigue | Does not interfere with daily activities | | Some interference with activity | Significant; prevents daily  activity | Visit to the emergency room*  OR  Hospitalization |
| Myalgia | Does not interfere with daily activities | | Some interference with activity | Significant; prevents daily  activity | Visit to the emergency room*  OR  Hospitalization |
| Chills | Feeling slightly cold; chills, teeth chattering | | Moderate whole-body shivering, requires use of opioids | Severe or prolonged, does not respond to opioids | ----- |
| Anorexia | Loss of appetite without alteration in eating habits | | Oral intake altered without significant weight loss or malnutrition; oral nutritional supplements indicated | Associated with significant weight loss or malnutrition (e.g., inadequate oral caloric and/or fluid intake); tube feeding, or Total parenteral nutrition indicated | ----- |
| Cough | Mild symptoms; nonprescription intervention indicated | | Moderate symptoms, medical intervention indicated; limiting instrumental activities of daily living | Severe symptoms; limiting self-care activities of daily living | ----- |
| Arthralgia | Mild pain | | Moderate pain; limiting instrumental activities of daily living | Severe pain; limiting self-care activities of daily living | ----- |
| Pruritus | Mild or localized; topical intervention indicated | | Widespread and intermittent; skin changes from scratching (e.g., edema, papulation, excoriations, lichenification, oozing/crusts); oral intervention indicated; limiting instrumental activities of daily living | Widespread and constant; limiting self-care activities of daily living or sleep; systemic corticosteroid or immunosuppressive therapy indicated | ----- |
| Skin  rash (exanthema)^†^ | Present, but asymptomatic | | Symptomatic (pruritus/pain), but interferes only slightly with daily activities | Symptomatic, prevents daily activities | Visit to the emergency room*  OR  Hospitalization |
| Allergic reaction | Systemic intervention not indicated | | Oral intervention indicated | Bronchospasm; hospitalization indicated for clinical sequelae; intravenous intervention indicated | Life-threatening consequences; urgent intervention indicated |

| **Table S3. Severity grading criteria for unsolicited adverse events*.*** | |
| --- | --- |
| **GRADE 1 (Mild)** | Transient (<48 hours) or mild discomfort; no medical intervention/therapy required |
| **GRADE 2 (Moderate)** | Mild to moderate limitation in activity - some assistance may be needed; no or minimal medical intervention/therapy required |
| **GRADE 3 (Severe)** | Marked limitation in activity, some assistance usually required; medical intervention/therapy required, hospitalizations possible |
| **GRADE 4**  **(Life-threatening)** | Extreme limitation in activity, significant assistance required; significant medical intervention/therapy required, hospitalization or hospice care probable |
| **GRADE 5** | Death |

| **Table S4. Solicited Local and Systemic Adverse Reactions between Participants in the 18-59 and ≥60 years old group, Classified by Severity Grade*** | | | | | | | | | | | | | |
| --- | --- | --- | --- | --- | --- | --- | --- | --- | --- | --- | --- | --- | --- |
| **Adverse Reaction**** | | **18-59 years** | | | | |  | **≥ 60 years** | | | | | **p-value ***** |
|  |  | **(n = 397)** | | | | |  | **(n = 37)** | | | | |  |
|  |  | **Grade 1** | **Grade 2** | **Grade 3** | **Grade 4** | **Total** |  | **Grade 1** | **Grade 2** | **Grade 3** | **Grade 4** | **Total** |  |
| **Local reactions** | |  |  |  |  |  |  |  |  |  |  |  |  |
| Pain, n (%) | | 183 (46.1) | 3 (0.8) | 1 (0.3) | 0 (0.0) | 187(47.1) |  | 10 (27.0) | 1 (2.7) | 0 (0.0) | 0 (0.0) | 11 (29.7) | **0.009** |
|  | Placebo | 45 (29.6) | 2 (1.3) | 0 (0.0) | 0 (0.0) | 47 (30.9) |  | 3 (25.0) | 0 (0.0) | 0 (0.0) | 0 (0.0) | 3 (25.0) | 0.466 |
|  | Vaccine | 138 (56.3) | 1 (0.4) | 1 (0.4) | 0 (0.0) | 140(57.1) |  | 7 (28.0) | 1 (4.0) | 0 (0.0) | 0 (0.0) | 8 (32.0) | **0.012** |
| Pruritus, n (%) | | 23 (5.8) | 0 (0.0) | 0 (0.0) | 0 (0.0) | 23 (5.8) |  | 0 (0.0) | 0 (0.0) | 0 (0.0) | 0 (0.0) | 0 (0.0) | 0.244 |
|  | Placebo | 4 (2.6) | 0 (0.0) | 0 (0.0) | 0 (0.0) | 4 (2.6) |  | 0 (0.0) | 0 (0.0) | 0 (0.0) | 0 (0.0) | 0 (0.0) | 0.999 |
|  | Vaccine | 19 (7.8) | 0 (0.0) | 0 (0.0) | 0 (0.0) | 19 (7.8) |  | 0 (0.0) | 0 (0.0) | 0 (0.0) | 0 (0.0) | 0 (0.0) | 0.229 |
| Induration, n (%) | | 17 (4.3) | 1 (0.3) | 1 (0.3) | 0 (0.0) | 19 (4.9) |  | 3 (8.1) | 0 (0.0) | 0 (0.0) | 0 (0.0) | 3 (8.1) | 0.466 |
|  | Placebo | 1 (0.7) | 0 (0.0) | 0 (0.0) | 0 (0.0) | 1 (0.7) |  | 0 (0.0) | 0 (0.0) | 0 (0.0) | 0 (0.0) | 0 (0.0) | --- |
|  | Vaccine | 16 (6.5) | 1 (0.4) | 1 (0.4) | 0 (0.0) | 18 (7.3) |  | 3 (12.0) | 0 (0.0) | 0 (0.0) | 0 (0.0) | 0 (0.0) | 0.468 |
| Erythema, n (%) | | 15 (3.8) | 1 (0.3) | 0 (0.0) | 0 (0.0) | 16 (4.1) |  | 1 (2.7) | 0 (0.0) | 0 (0.0) | 0 (0.0) | 1 (2.7) | 0.999 |
|  | Placebo | 3 (2.0) | 0 (0.0) | 0 (0.0) | 0 (0.0) | 3 (2.0) |  | 1 (8.3) | 0 (0.0) | 0 (0.0) | 0 (0.0) | 1 (8.3) | 0.191 |
|  | Vaccine | 12 (4.9) | 1 (0.4) | 0 (0.0) | 0 (0.0) | 13 (5.3) |  | 0 (0.0) | 0 (0.0) | 0 (0.0) | 0 (0.0) | 0 (0.0) | 0.605 |
| Swelling, n (%) | | 9 (2.3) | 3 (0.8) | 0 (0.0) | 0 (0.0) | 12 (3.1) |  | 1 (2.7) | 0 (0.0) | 0 (0.0) | 0 (0.0) | 1 (2.7) | 0.999 |
|  | Placebo | 3 (2.0) | 1 (0.7) | 0 (0.0) | 0 (0.0) | 4 (2.7) |  | 0 (0.0) | 0 (0.0) | 0 (0.0) | 0 (0.0) | 0 (0.0) | 0.999 |
|  | Vaccine | 6 (2.4) | 2 (0.8) | 0 (0.0) | 0 (0.0) | 8 (3.2) |  | 1 (4.0) | 0 (0.0) | 0 (0.0) | 0 (0.0) | 1 (4.0) | 0.999 |
| **Systemic reactions** | |  |  |  |  |  |  |  |  |  |  |  |  |
| Headache, n (%) | | 86 (21.7) | 77 (19.4) | 3 (0.8) | 2 (0.5) | 168(42.4) |  | 5 (13.5) | 5 (13.5) | 0 (0.0) | 0 (0.0) | 10 (27.0) | **0.026** |
|  | Placebo | 25 (16.4) | 27 (17.8) | 0 (0.0) | 1 (0.7) | 53 (34.9) |  | 2 (16.7) | 1 (8.3) | 0 (0.0) | 0 (0.0) | 3 (25.0) | 0.713 |
|  | Vaccine | 61 (24.9) | 50 (20.4) | 3 (1.2) | 1 (0.4) | 115(46.9) |  | 3 (12.0) | 4 (16.0) | 0 (0.0) | 0 (0.0) | 7 (28.0) | **0.03** |
| Fatigue, n (%) | | 67 (16.9) | 31 (7.8) | 2 (0.5) | 1 (0.3) | 101 (25.4) |  | 3 (8.1) | 3 (8.1) | 1 (2.7) | 0 (0.0) | 7 (18.9) | 0.23 |
|  | Placebo | 19 (12.5) | 13 (8.6) | 1 (0.7) | 1 (0.7) | 34 (32.4) |  | 2 (16.7) | 0 (0.0) | 1 (8.3) | 0 (0.0) | 3 (25.0) | 0.999 |
|  | Vaccine | 48 (19.6) | 18 (7.3) | 1 (0.4) | 0 (0.0) | 67 (27.3) |  | 1 (4.0) | 3 (12.0) | 0 (0.0) | 0 (0.0) | 4 (16.0) | 0.198 |
| Myalgia, n (%) | | 60 (15.1) | 21 (5.3) | 2 (0.5) | 0 (0.0) | 83 (20.9) |  | 4 (10.8) | 1 (2.7) | 1 (2.7) | 0 (0.0) | 6 (16.2) | 0.264 |
|  | Placebo | 18 (11.8) | 8 (5.3) | 1 (0.7) | 0 (0.0) | 27 (17.8) |  | 1 (8.3) | 0 (0.0) | 1 (8.3) | 0 (0.0) | 2 (16.7) | 0.627 |
|  | Vaccine | 42 (17.1) | 13 (5.3) | 1 (0.4) | 0 (0.0) | 56 (22.8) |  | 3 (12.0) | 1 (4.0) | 0 (0.0) | 0 (0.0) | 4 (16.0) | 0.405 |
| Diarrhea, n (%) | | 56 (14.1) | 11 (2.8) | 0 (0.0) | 0 (0.0) | 67 (16.9) |  | 4 (10.8) | 0 (0.0) | 0 (0.0) | 0 (0.0) | 4 (10.8) | 0.142 |
|  | Placebo | 15 (9.9) | 5 (3.3) | 0 (0.0) | 0 (0.0) | 20 (13.2) |  | 2 (16.7) | 0 (0.0) | 0 (0.0) | 0 (0.0) | 2 (16.7) | 0.999 |
|  | Vaccine | 41 (16.7) | 6 (2.4) | 0 (0.0) | 0 (0.0) | 47 (19.1) |  | 2 (8.0) | 0 (0.0) | 0 (0.0) | 0 (0.0) | 2 (8.0) | 0.272 |
| Nausea, n (%) | | 39 (9.8) | 5 (1.3) | 2 (0.5) | 0 (0.0) | 46 (11.6) |  | 3 (8.1) | 1 (2.7) | 0 (0.0) | 0 (0.0) | 4 (10.8) | 0.999 |
|  | Placebo | 15 (9.9) | 3 (2.0) | 0 (0.0) | 0 (0.0) | 18 (11.9) |  | 1 (8.3) | 0 (0.0) | 0 (0.0) | 0 (0.0) | 1 (8.3) | 0.999 |
|  | Vaccine | 24 (9.8) | 2 (0.8) | 2 (0.8) | 0 (0.0) | 28 (11.4) |  | 2 (8.0) | 1 (4.0) | 0 (0.0) | 0 (0.0) | 3 (12.0) | 0.999 |
| Arthralgia, n (%) | | 21 (5.3) | 7 (1.8) | 1 (0.3) | 0 (0.0) | 29 (7.3) |  | 1 (2.7) | 1 (2.7) | 0 (0.0) | 0 (0.0) | 2 (5.4) | 0.999 |
|  | Placebo | 8 (5.3) | 3 (2.0) | 0 (0.0) | 0 (0.0) | 10 (7.3) |  | 0 (0.0) | 0 (0.0) | 0 (0.0) | 0 (0.0) | 0 (0.0) | 0.999 |
|  | Vaccine | 13 (5.3) | 4 (1.6) | 1 (0.4) | 0 (0.0) | 18 (7.3) |  | 1 (4.0) | 1 (4.0) | 0 (0.0) | 0 (0.0) | 2 (8.0) | 0.999 |
| Anorexia, n (%) | | 20 (5.0) | 9 (2.3) | 1 (0.3) | 0 (0.0) | 30 (7.6) |  | 2 (5.4) | 1 (2.7) | 0 (0.0) | 0 (0.0) | 3 (8.1) | 0.486 |
|  | Placebo | 7 (4.6) | 4 (2.6) | 0 (0.0) | 0 (0.0) | 11 (7.2) |  | 0 (0.0) | 1 (8.3) | 0 (0.0) | 0 (0.0) | 1 (8.3) | 0.48 |
|  | Vaccine | 13 (5.3) | 5 (2.0) | 1 (0.4) | 0 (0.0) | 19 (7.7) |  | 2 (8.0) | 0 (0.0) | 0 (0.0) | 0 (0.0) | 2 (8.0) | 0.677 |
| Pruritus, n (%) | | 13 (3.3) | 2 (0.5) | 0 (0.0) | 0 (0.0) | 15 (3.8) |  | 1 (2.7) | 0 (0.0) | 0 (0.0) | 0 (0.0) | 1 (2.7) | 0.999 |
|  | Placebo | 2 (1.3) | 0 (0.0) | 0 (0.0) | 0 (0.0) | 2 (1.3) |  | 0 (0.0) | 0 (0.0) | 0 (0.0) | 0 (0.0) | 0 (0.0) | 0.999 |
|  | Vaccine | 11 (4.5) | 2 (0.8) | 0 (0.0) | 0 (0.0) | 13 (5.3) |  | 1 (4.0) | 0 (0.0) | 0 (0.0) | 0 (0.0) | 1 (4.0) | 0.999 |
| Exanthema, n (%) | | 6 (1.5) | 3 (0.8) | 0 (0.0) | 0 (0.0) | 9 (2.3) |  | 0 (0.0) | 0 (0.0) | 0 (0.0) | 0 (0.0) | 0 (0.0) | 0.605 |
|  | Placebo | 1 (0.7) | 0 (0.0) | 0 (0.0) | 0 (0.0) | 1 (0.7) |  | 0 (0.0) | 0 (0.0) | 0 (0.0) | 0 (0.0) | 0 (0.0) | 0.999 |
|  | Vaccine | 5 (2.0) | 3 (1.2) | 0 (0.0) | 0 (0.0) | 8 (3.2) |  | 0 (0.0) | 0 (0.0) | 0 (0.0) | 0 (0.0) | 0 (0.0) | 0.999 |
| Allergy, n (%) | | 5 (1.3) | 2 (0.5) | 0 (0.0) | 0 (0.0) | 7 (1.8) |  | 0 (0.0) | 2 (5.4) | 0 (0.0) | 0 (0.0) | 2 (5.4) | 0.587 |
|  | Placebo | 0 (0.0) | 0 (0.0) | 0 (0.0) | 0 (0.0) | 0 (0.0) |  | 0 (0.0) | 1 (8.3) | 0 (0.0) | 0 (0.0) | 1 (8.3) | --- |
|  | Vaccine | 5 (2.0) | 2 (0.8) | 0 (0.0) | 0 (0.0) | 7 (2.8) |  | 0 (0.0) | 1 (4.0) | 0 (0.0) | 0 (0.0) | 1 (4.0) | 0.593 |
| Vomiting, n (%) | | 5 (1.3) | 2 (0.5) | 1 (0.3) | 0 (0.0) | 8 (2.1) |  | 0 (0.0) | 0 (0.0) | 0 (0.0) | 0 (0.0) | 0 (0.0) | 0.999 |
|  | Placebo | 3 (2.0) | 0 (0.0) | 0 (0.0) | 0 (0.0) | 3 (2.0) |  | 0 (0.0) | 0 (0.0) | 0 (0.0) | 0 (0.0) | 0 (0.0) | --- |
|  | Vaccine | 2 (0.8) | 2 (0.8) | 1 (0.4) | 0 (0.0) | 5 (2.0) |  | 0 (0.0) | 0 (0.0) | 0 (0.0) | 0 (0.0) | 0 (0.0) | 0.999 |
| Fever (>37.8ºC), n (%) | | 0 (0.0) | 2 (0.5) | 0 (0.0) | 0 (0.0) | 2 (0.5) |  | 0 (0.0) | 0 (0.0) | 0 (0.0) | 0 (0.0) | 0 (0.0) | 0.999 |
|  | Placebo | 0 (0.0) | 1 (0.7) | 0 (0.0) | 0 (0.0) | 1 (0.7) |  | 0 (0.0) | 0 (0.0) | 0 (0.0) | 0 (0.0) | 0 (0.0) | 0.999 |
|  | Vaccine | 0 (0.0) | 1 (0.4) | 0 (0.0) | 0 (0.0) | 1 (0.4) |  | 0 (0.0) | 0 (0.0) | 0 (0.0) | 0 (0.0) | 0 (0.0) | 0.999 |
| *Percentages were calculated from the total number of participants in each group.  **Data in the table were reported within 14 days post any of the two doses.  ***Comparison of having or not the reaction, between age group. | | | | | | | | | | | | | |

| Table S5. Evaluation of the immune response of participants seropositive at enrollment and breakthrough cases. | | | | | | | | | | | | | | | | | | | | | | |
| --- | --- | --- | --- | --- | --- | --- | --- | --- | --- | --- | --- | --- | --- | --- | --- | --- | --- | --- | --- | --- | --- | --- |
| Antibodies detection (GMT) | | **Day 0** | | | | | |  | **Day 28** | | | | | | |  | **Day 42** | | | | | |
|  |  | **Seropositive** | | | | **Breakthrough** | |  | **Seropositive** | | | | **Breakthrough** | | |  | **Seropositive** | | | | **Breakthrough** | |
| Anti-N | | 1 | 800 | 1 | 800 | 1 | 1 |  | 800 | 12800 | 3200 | 6400 | | 50 | 1 |  | 400 | 1600 | - | 6400 | 1 | 1 |
| Anti-S1-RBD | | 1 | 6400 | 800 | 6400 | 1 | 1 |  | 12800 | 12800 | 3200 | 6400 | | 6400 | 200 |  | 3200 | 6400 | 1600 | 6400 | 6400 | 1600 |
| Neutralizing Ab | | 64 | 64 | 1 | - | 1 | 1 |  | 64 | 128 | 128 | - | | 64 | 4 |  | - | - | - | - | - | - |
| Fold change ELISPOT (Spot forming cells) | | **Seropositive** | | | | **Breakthrough** | |  | **Seropositive** | | | | **Breakthrough** | | |  | **Seropositive** | | | | **Breakthrough** | |
| MP-R | IFN-γ | 1.00 | 1.00 | 1.00 | 1.00 | 1.00 | 1.00 |  | 0.09 | 0.10 | 3.00 | 4.59 | | 1.29 | 2.02 |  | 0.82 | 0.18 | 7.04 | 3.18 | 0.49 | 0.01 |
|  | IL-4 | 1.00 | 1.00 | 1.00 | 1.00 | 1.00 | 1.00 |  | 0.50 | 1.00 | 1.00 | 1.00 | | 1.00 | 1.00 |  | 1.00 | 1.00 | 1.00 | 1.00 | 1.00 | 1.00 |
| MP-S | IFN-γ | 1.00 | 1.00 | 1.00 | 1.00 | 1.00 | 1.00 |  | 0.07 | 0.43 | 4.18 | 0.30 | | 2.64 | 0.07 |  | 0.75 | 0.16 | 4.53 | 0.18 | 2.45 | 0.01 |
|  | IL-4 | 1.00 | 1.00 | 1.00 | 1.00 | 1.00 | 1.00 |  | 1.00 | 1.00 | 1.00 | 1.00 | | 1.00 | 1.00 |  | 3.00 | 1.00 | 1.00 | 1.00 | 1.00 | 1.00 |
| Fold change Flow Cytometry (# of cells) | | **Seropositive** | | | | **Breakthrough** | |  | **Seropositive** | | | | **Breakthrough** | | |  | **Seropositive** | | | | **Breakthrough** | |
| MP-R | CD4+CD69+ | 1.00 | 1.00 | 1.00 | 1.00 | 1.00 | 1.00 |  | 0.81 | 0.89 | 4.00 | 1.45 | | 0.00 | 1.31 |  | 0.90 | 0.82 | 2.50 | 1.60 | 0.00 | 0.34 |
|  | CD4+CD69+ IFN-γ+ | 1.00 | 1.00 | 1.00 | 1.00 | 1.00 | 1.00 |  | 1.50 | 1.00 | 0.00 | 5.00 | | 0.00 | 4.00 |  | 1.67 | 0.00 | 1.00 | 5.00 | 0.00 | 1.00 |
|  | CD4+CD69+ IL-2+ | 1.00 | 1.00 | 1.00 | 1.00 | 1.00 | 1.00 |  | 0.00 | 0.67 | 3.00 | 1.83 | | 1.00 | 0.80 |  | 2.00 | 0.33 | 3.00 | 2.00 | 1.00 | 0.00 |
|  | CD4+CD69+ TNF+ | 1.00 | 1.00 | 1.00 | 1.00 | 1.00 | 1.00 |  | 0.36 | 1.00 | 0.00 | 10.00 | | 1.00 | 4.00 |  | 0.55 | 2.00 | 0.00 | 3.00 | 1.00 | 0.00 |
| MP-S | CD4+CD69+ | 1.00 | 1.00 | 1.00 | 1.00 | 1.00 | 1.00 |  | 0.51 | 0.26 | 2.25 | 0.22 | | 4.97 | 0.82 |  | 0.67 | 0.93 | 1.75 | 0.81 | 3.06 | 0.51 |
|  | CD4+CD69+ IFN-γ+ | 1.00 | 1.00 | 1.00 | 1.00 | 1.00 | 1.00 |  | 0.43 | 0.50 | 0.00 | 0.15 | | 0.50 | 1.50 |  | 0.29 | 1.00 | 0.00 | 0.85 | 0.50 | 0.50 |
|  | CD4+CD69+ IL-2+ | 1.00 | 1.00 | 1.00 | 1.00 | 1.00 | 1.00 |  | 0.00 | 0.00 | 2.50 | 0.80 | | 2.00 | 1.33 |  | 0.88 | 2.00 | 0.50 | 0.40 | 1.00 | 0.67 |
|  | CD4+CD69+ TNF+ | 1.00 | 1.00 | 1.00 | 1.00 | 1.00 | 1.00 |  | 0.50 | 0.50 | 0.00 | 0.20 | | 1.00 | 2.00 |  | 0.75 | 3.50 | 0.00 | 0.90 | 0.00 | 1.00 |

| Table S6. Fold-change analysis for Flow Cytometry assays in vaccinated participants, aged 18-59 or ≥60 years old | | | | | | | | | |
| --- | --- | --- | --- | --- | --- | --- | --- | --- | --- |
|  | |  | **18-59** | | |  | **≥60** | | |
| Mega Pool Parameter | |  | **Day 0** | **Day 28** | **Day 42** |  | **Day 0** | **Day 28** | **Day 42** |
| MP-S | CD4+CD69+ |  | 1,00 | 2.17 (0.3) | 1.67 (0.37) |  | 1,00 | 1.97 (0.45) | 3.24 (2.72) |
|  | CD4+CD69+ IFN-𝛾+ |  | 1,00 | 2.2 (0.45) | 1.64 (0.37) |  | 1,00 | 0.76 (0.41) | 1.04 (0.65) |
|  | CD4+CD69+ IL-2+ |  | 1,00 | 2.26 (0.59) | 1.62 (0.44) |  | 1,00 | 0.43 (0.2) | 0.44 (0.34) |
|  | CD4+CD69+ TNF+ |  | 1,00 | 1.69 (0.59) | 1.6 (0.97) |  | 1,00 | 0.29 (0.18) | 0.15 (0.11) |
|  | CD8+CD69+ |  | 1,00 | 2.11 (0.45) | 1.66 (0.36) |  | 1,00 | 1.09 (0.37) | 1.92 (0.94) |
|  | CD8+CD69+ IFN-𝛾+ |  | 1,00 | 2.5  (0.82) | 1.85 (0.92) |  | 1,00 | 0.32 (0.15) | 1.97 (1.52) |
|  | CD8+CD69+ IL-2+ |  | 1,00 | 1.38 (0.52) | 1.59 (0.74) |  | 1,00 | 0.57 (0.43) | 1.28 (0.93) |
|  | CD8+CD69+ TNF+ |  | 1,00 | 0.82 (0.31) | 0.96 (0.32) |  | 1,00 | 0 (0) | 0.33 (0.33) |
| MP-R | CD4+CD69+ |  | 1,00 | 2.2  (0.39) | 1.28 (0.21) |  | 1,00 | 1.42 (0.5) | 2.83 (1.25) |
|  | CD4+CD69+ IFN-𝛾+ |  | 1,00 | 2.26 (0.61) | 1.2 (0.3) |  | 1,00 | 1.07 (0.6) | 0.56 (0.29) |
|  | CD4+CD69+ IL-2+ |  | 1,00 | 1.4  (0.22) | 0.95 (0.2) |  | 1,00 | 1.5 (0.66) | 0.81 (0.3) |
|  | CD4+CD69+ TNF+ |  | 1,00 | 1.51  (0.5) | 0.5 (0.17) |  | 1,00 | 0.86 (0.26) | 1.88 (1.6) |
|  | CD8+CD69+ |  | 1,00 | 1.64 (0.27) | 1.39 (0.48) |  | 1,00 | 1.56 (0.49) | 1.66 (0.57) |
|  | CD8+CD69+ IFN-𝛾+ |  | 1,00 | 1.59 (0.52) | 2.05 (0.91) |  | 1,00 | 0.36 (0.24) | 1.44 (0.95) |
|  | CD8+CD69+ IL-2+ |  | 1,00 | 0.9  (0.26) | 1.07 (0.33) |  | 1,00 | 0.29 (0.18) | 1.63 (1.12) |
|  | CD8+CD69+ TNF+ |  | 1,00 | 0.56 (0.14) | 0.8 (0.3) |  | 1,00 | 0 (0) | 0.13 (0.13) |
| MP-CD8A | CD4+CD69+ |  | 1,00 | 1.09 (0.16) | 0.76 (0.18) |  | 1,00 | 1.57 (0.38) | 1.13 (0.4) |
|  | CD4+CD69+ IFN-𝛾+ |  | 1,00 | 1.1  (0.46) | 0.41 (0.1) |  | 1,00 | 0.68 (0.34) | 0.79 (0.41) |
|  | CD4+CD69+ IL-2+ |  | 1,00 | 0.21 (0.14) | 0.58 (0.26) |  | 1,00 | 2.01 (1.02) | 1.26 (0.49) |
|  | CD4+CD69+ TNF+ |  | 1,00 | 0.77 (0.38) | 1.33 (0.45) |  | 1,00 | 1.87 (0.62) | 0.71 (0.14) |
|  | CD8+CD69+ |  | 1,00 | 1.35 (0.17) | 0.85 (0.25) |  | 1,00 | 2.37 (0.64) | 0.91 (0.17) |
|  | CD8+CD69+ IFN-𝛾+ |  | 1,00 | 1.08 (0.17) | 0.53 (0.18) |  | 1,00 | 3.7 (1.42) | 1.5 (0.63) |
|  | CD8+CD69+ IL-2+ |  | 1,00 | 1.39 (0.59) | 0.35 (0.19) |  | 1,00 | 1.16 (0.39) | 0.47 (0.19) |
|  | CD8+CD69+ TNF+ |  | 1,00 | 0.88  (0.4) | 0 (0) |  | 1,00 | 0.14 (0.14) | 0.29 (0.18) |
| MP-CD8B | CD4+CD69+ |  | 1,00 | 1.41 (0.38) | 0.88 (0.26) |  | 1,00 | 1.54 (0.36) | 1.11 (0.43) |
|  | CD4+CD69+ IFN-𝛾+ |  | 1,00 | 1.96 (0.68) | 0.83 (0.31) |  | 1,00 | 0.61 (0.41) | 0.7 (0.34) |
|  | CD4+CD69+ IL-2+ |  | 1,00 | 0.33 (0.21) | 0.57 (0.43) |  | 1,00 | 2 (0.83) | 1.18 (0.35) |
|  | CD4+CD69+ TNF+ |  | 1,00 | 1.43 (0.44) | 1.93 (1.25) |  | 1,00 | 1.7 (0.51) | 1.83 (0.89) |
|  | CD8+CD69+ |  | 1,00 | 2.68  (1.1) | 1.04 (0.4) |  | 1,00 | 1.86 (0.36) | 1.35 (0.39) |
|  | CD8+CD69+ IFN-𝛾+ |  | 1,00 | 2.75 (1.06) | 1.06 (0.4) |  | 1,00 | 1.49 (0.5) | 0.92 (0.34) |
|  | CD8+CD69+ IL-2+ |  | 1,00 | 3  (1.23) | 0.57 (0.43) |  | 1,00 | 1.26 (0.48) | 1.43 (0.61) |
|  | CD8+CD69+ TNF+ |  | 1,00 | 0.57  (0.2) | 0.57 (0.43) |  | 1,00 | 0 (0) | 0.86 (0.34) |
| Values represent the mean (SD) | | | | | | | | | |

**Supplementary Information: Materials and Methods**

**Study design, randomization, and participants**

This clinical trial (clinicaltrials.gov NCT04651790) was conducted in Chile at eight different sites, six located in Santiago city (Metropolitan Region) and two in the V Region of Valparaiso. This trial was approved by each Institutional Ethical Committee and the Chilean Public Health Institute (ISP Chile, number Nº 24204/20). Participants did not receive any payment for their participation. As vaccine and placebo pre-loaded syringes were different in appearance, a non-blind team of nurses was in charge of the product and its administration. This nurse team did not participate in any other study procedure.

**Vaccine composition**

CoronaVac consists of 3 µg of β-propiolactone inactivated SARS-CoV-2 (strain CZ02) with aluminum hydroxide as an adjuvant in 0.5 mL ^9^. Sodium chloride, monosodium hydrogen phosphate, and disodium hydrogen phosphate are excipients, and water for injection is included as solvent. Placebo consists of 0.5 mL of aqueous suspension for injection with aluminum hydroxide and excipients.

**Evaluation of SARS-CoV-2 N protein in vaccine doses**

Vials of placebo (Lot# 2020041602) and vaccine (Lot #20200412) (37.5 µL) and 2 µg of SARS-CoV-2 denatured recombinant N protein (1 μg, SinoBiological, 40588-V08B) were resolved in 12% SDS-PAGE. Gels were transferred to nitrocellulose membranes and incubated with 5% BSA (BM-0150, Winkler, RM, Chile) – PBS - 0.05% Tween-20 to block nonspecific binding for 2h at room temperature or O.N. at 4ºC. Membranes were incubated with a mouse IgG anti-N-SARS-CoV-2 (Sinobiological, Code 40143-MM05, Lot Nº HB14FE2110) at a 1:1,000 dilution in 3% BSA – PBS - 0.05% Tween-20, O.N. at 4ºC. Then, membranes were washed 3 times for 5 min in PBS - 0.05% Tween-20 and incubated with an anti-mouse IgG-HRP-conjugated antibody (Invitrogen, Code 62-6520, Lot Nº UJ287702) for 1h at RT. Membranes were washed 3 times for 5 min in PBS - 0.05% Tween-20 and then twice with PBS for 5 min. The reaction was revealed using PierceTM ECL Western Blotting substrate (32106, ThermoFisher, USA) and images were acquired in Invitrogen™ iBright™ CL1500 Imaging System.

**Procedures**

Prior to vaccination, a urine test was performed on all female participants to rule out pregnancy, which is an exclusion criterion. Also, blood samples and nasopharyngeal swabs were obtained for all participants to evaluate past or ongoing SARS-CoV-2 infection. After vaccination, participants were then kept in observation for 60 min, to evaluate possible immediate adverse events (AEs). Participants were then instructed to record solicited and unsolicited AEs, SAEs, Events of Special Interest, medications, and COVID-19 symptoms through an eCRF, with daily reminders sent to them the first 42 days.

For sera samples, 20 mL of blood were collected in tubes without anticoagulant and distributed in 2 tubes of 10 mL per participant (BD Vacutainer Clot Activator tubes #367896). After collection, blood was allowed to clot for at least 1h at RT. Samples were then centrifuged in a refrigerated centrifuge with a horizontal rotor at 1,300 x *g* for 10 min at 22ºC. Then, serum was carefully collected and transferred into clean polypropylene cryotubes for freezing and stored at –80°C until use. Hemolyzed samples were rejected.

For PBMC isolation, whole blood was collected in 3 heparinized tubes (BD Vacutainer #367874, 10 mL) and stored at RT until processing. Whole blood was then diluted with PBS (1:1) and centrifuged for 10 min at 1,200 x g (RT) in SepMate™ tubes (StemCell Technologies) with density-gradient medium (Lymphoprep) to separate the cellular fraction and plasma. The plasma was then carefully removed from the upper layer, and PBMCs were isolated by pouring them into a fresh tube. Isolated PBMC were washed twice with PBS, counted in an automated cell counter (Logos Biosystems #L40001), and cryopreserved in FBS (Industrial Biologicals) supplemented with 10% DMSO (Chem Cruz). All PBMC samples were stored in liquid nitrogen until use.

To assess the presence of anti-SARS-CoV-2 antibodies, blood samples from 39 participants obtained at days 0 (baseline), 14, 28, and 42 after the first dose were analyzed. The quantitative measurement of human IgG antibodies against the RBD domain of the S1 protein (S1-RBD) and against the N protein of SARS-CoV-2 was determined using the RayBio COVID-19 (SARS-CoV-2) Human Antibody Detection Kit (Indirect ELISA method) (Cat #IEQ-CoVS1RBD-IgG & #IEQ-CovN-IgG). Briefly, these kits consist of 96-well plates coated with the S1-RBD protein segment or the N protein of SARS-CoV-2. Sera samples were serially diluted starting at a 50-fold dilution until a 6,400-fold dilution. After 1h of incubation at RT, the plates were washed, and a biotinylated anti-human IgG antibody provided in the kit was added and incubated for 30 min at RT. Plates were washed, and then an HRP-conjugated streptavidin was added and incubated for 30 min at RT. Plates were rewashed, and the TMB substrate solution supplied in the kit was added. Finally, a stop solution provided in the kit was added, and absorbance was measured at 450 nm in an ELISA plate reader (Biotek, Ref. 1506021). As controls, dilutions of the First WHO International Standard for anti-SARS-CoV-2 immunoglobulin (human - NIBSC code: 20/13) were included. Additional controls included samples of participants seropositive or seronegative at recruitment or inoculated with placebo. To calculate the end titers, the cut-off value for each test was determined as ≥2.1 times the average OD_450nm_ value of a panel of 27-29 seronegative serum samples. These serum samples were obtained before vaccination from seronegative participants. Seropositivity was determined as the highest dilution that reached >2.1 times the OD_450nm_ cut-off value. Seroconversion was defined as an increase of at least four times the titer at baseline.

The neutralizing capacities of the antibodies in the samples of the participants were evaluated through the SARS-CoV-2 sVNT kit from Genscript (cat. number L00847-A). Assays were performed according to the instructions of the manufacturer. Briefly, serial dilutions of the serum were prepared and then incubated with the HRP-RBD reagent supplied in the kit for 30 min at 37ºC to allow the binding of neutralizing antibodies to S1-RBD. Sera and controls previously incubated with the HRP-RBD were added to the ELISA plate pre-coated with the human angiotensin-converting enzyme 2 (hACE2) protein and incubated for 15 min at 37ºC. After the incubation, samples were discarded, and plates were washed. Finally, a developing solution provided in the kit was added for 15 min at RT and then quenched with a stop solution also supplied. Plates were read at 450 nm in a microplate reader (Biotek, Ref. 1506021). The inhibition rate of HRP-RBD binding was calculated as follows: [OD_450nm_ value of negative control -OD_450nm_ value of sample]/OD450 of negative control*100]. To calculate the inhibition rate, the average OD_450nm_ from a panel of negative serum samples was used. A cut-off >30% was used to define if samples were positive or not for the presence of anti-SARS-CoV2 neutralizing antibodies. Seroconversion rates were defined as the change from seronegative at baseline to seropositive. The methods used in this study for the detection of total and neutralizing antibodies were validated with the First WHO International Standard Anti-SARS-CoV-2 Immunoglobulin (Cat# 20/136 and #20/268), kindly provided by PATH.

To assess the cellular immune response, ELISPOT and flow cytometry assays were performed using PBMCs from 36 participants. Upon thawing, cells were resuspended in fresh media in a 1:10 dilution to remove DMSO remnants from the freezing media. Then, cells were centrifuged, resuspended in fresh media, and counted in an automated cell counter (Logos Biosystems #L40001). Cells were adjusted to 6x10^6^ cells/mL and kept at 37ºC, 5% CO_2_ for 15 min until use in the corresponding assay. ELISPOT plates containing a PVDF membrane were activated with 15 µL of 70% ethanol (Merck), washed three times with sterile 1x PBS, and then coated with human IFN-γ and IL-4 capture antibodies (1:250 and 1:125, respectively, CTL). After 3h of activation at RT, plates were washed two times with PBS and two times with PBS-Tween 20 0.05%. Stimulus included in these assays considers the use of Mega Pools (MPs) of peptides derived from SARS-CoV-2 proteins, previously described ^13^. Two MPs composed of peptides from the S protein (MP-S) and the remaining proteins of the viral particle (MP-R) were used, as previously described ^13^. These peptides were determined *in silico* to optimally stimulate CD4^+^ T cell. Also, two MPs composed of peptides from the proteome of SARS-CoV-2 (CD8-A and CD8-B) were used, as previously described ^13^. These peptides were determined *in silico* to optimally stimulate CD8^+^ T cell. As positive controls, an independent stimulation performed with 5 mg/mL of Concanavalin A (ConA) (Sigma Life Science #C5275-5MG) and with an MP of peptides derived from cytomegalovirus proteins (MP-CMV) for the stimulation of both CD4^+^ and CD8^+^ T cells ^13^. As a vehicle control, DMSO 1% (Merck #317275) was included. A total of 3x10^5^ cells in 50 µL of media were added to each well containing 50 µL of media with the corresponding stimulus. The final concentration of each stimulus per well was 1 µg/mL (except for ConA and DMSO). Positive controls for ELISPOT assays considered 5x10^4^ cells/well instead of 3x10^5^ cells/well. For ELISPOT assays, cells were incubated for 48h at 37ºC, 5% CO_2_. After incubation, plates were washed 1 time with PBS and 3 times with PBS-Tween20. Then, anti-human IFN-γ (FITC) and anti-human IL-2 (Biotin) antibodies (1:1,000 and 1:1,000, respectively) were added and plates were incubated for 2h, RT. Plates were washed 3 more times with PBS-Tween 20 and then FITC-HRP and Streptavidin-AP (1:1,000) were added and plates were incubated for 1h, RT. After incubation, plates were washed 3 more times with PBS-Tween20. Then, plates were treated with the blue (15 min) and red (15 min) developer solution individually following the following the recommendations of the manufacturer. Plates were washed with tap water after each developer solution and allowed to dry for 24h prior to reading. To evaluate the number of T cells secreting IFN-γ, IL-4 or both, ELISPOT assays were performed with ImmunoSpot^®^ technology (ImmnunoSpot^®^ #hIFNgIL4-1M-10). Spot Forming Cells (SFCs) were counted on an ImmunoSpot^®^ S6 Micro Analyzer.

To characterize T cells and the production of cytokines by these populations, flow cytometry assays were performed 3x10^5^ cells per well were stimulated as described for the ELISPOT assays and after 24 h of incubation with the stimulus, samples were stained with several surface and intracellular markers. Antibodies used are as follow: for the extracellular staining, a mix of BD Horizon^TM^ Fixable Viability Stain 510 (BD Biosciences –  CAT 564406 – 1 µL per 1x10^6^ cells); anti-CD45RA BV421 (Biolegend - clone HI100); anti-CD14 V500 (BD Bioscience - clone M5E2); anti-CD19 V500 (BD Bioscience - clone HIB19); anti-CD3 AF-700 (Biolegend - clone OKT3); anti-CD69 PE (BD Bioscience - clone FN50); anti-CD8a BV-650 (BD Bioscience - clone RPA-T8); and anti-CD4 BV-605 (BD Bioscience - clone RPA-T4) was incubated for 45 min at 4ºC. Cells were washed twice with 200 µL of PEB buffer and then fixed and permeabilized with the BD Cytofix/Cytoperm (CAT 554714) kit, following the instructions of the manufacturer. Cells were then stained ON at 4ºC for intracellular cytokines using the following antibodies: anti-IL-2 PE-CF594 (BD Bioscience - clone 5344-111), anti-IFN-γ BV-786 (BD Bioscience - clone 4S.B3) and anti-TNF-ɑ PercP-Cy5.5 (BD Bioscience - clone MAb11). The next morning, cells were washed twice with 200 µL of PEB buffer and handed to the Flow Cytometry facility, for their acquisition by an LSRFortessa X-20 flow cytometer.

**Pseudotyped virus neutralization assay**

Anti-SARS-CoV-2 neutralizing antibodies were measured using an HIV-1 backbone expressing firefly luciferase as a reporter gene and pseudotyped with the SARS-CoV-2 spike glycoprotein (HIV-1-SΔ19) as previously described ^15^. Briefly, serum samples were initially diluted 1:4 in DMEM, serially diluted 1:3 up to 1:8,748 and then mixed with approximately 4.5 ng of p24 equivalents of HIV-1-SΔ19 in white 96-well plate. Plates were incubated during 1 hour at 37°C and then 100 mL of DMEM containing 1x10^4^ HEK-ACE2 cells was added to each well. Firefly luciferase activity was measured 48h later, using the Luciferase Assay System (Promega) in a Glomax®-96 microplate luminometer (Promega). Estimation of the ID50 was obtained using a 4-parameter nonlinear regression curve fit measured as the percent of neutralization determined by the difference in average relative light units (RLU) between test samples and pseudotyped virus controls as previously described ^15^. Data analyses and statistical analyses were carried out using GraphPad Prism v8.

**BioHermes surrogate virus neutralization assay**

The SARS-CoV-2 Neutralizing Antibodies Test (BSNAT) Kit from BioHermes (Ref. COV-S41) was used to detect neutralizing antibodies in serum against SARS-CoV-2. This kit is a blocking assay which simulates the neutralization process, based in the ELISA platform. The main components of the kit are an ELISA plate pre-coated with the human ACE2 (hACE2) protein and the SARS-CoV-2 S1-RBD fragment conjugated with horseradish peroxidase (HRP-RBD). Briefly, the first step consisted in preparing serial dilutions from serum samples with the sample dilutor provided in the kit. Then, samples and controls were incubated with the HRP-RBD during 10 min at 37ºC to allow the binding of neutralizing antibodies to RBD. Serums and controls previously incubated with HRP-RBD were added to ELISA plates (pre-coated with hACE2) and incubated for 20 min at 37ºC. After incubation, samples were discarded, and all the wells were washed five times. Finally, TMB solution was added and quenched after 15 minutes of incubation at room temperature. Plates were read at 450 nm in a microplate reader (EPOCH, Biotek ref. 1506021). The inhibition rate was calculated based on the negative control absorbances (negative control – sample/negative control*100), and 10% was considered as the cut-off value.

**Neutralization assays**

Vero E6 cells were infected with a SARS-CoV-2 strain obtained by viral isolation in tissue cultures (33782CL-SARS-CoV-2 strain). Neutralization assays were carried out by the reduction of cytopathic effect (CPE) in Vero E6 cells (ATCC CRL-1586). The titer of neutralizing antibodies was defined as the highest serum dilution that neutralized virus infection, at which the CPE was absent compared with the virus control wells (cells with CPE). Vero E6 cells (4×10^4^ cells/well) were seeded in 96-well plates. For neutralization assays, 100 µL of 33782CL-SARS-CoV-2 (at a dose of 100 TCID_50_) were incubated with serial dilutions of heat-inactivated sera samples (dilutions of 1:4, 1:8, 1:16, 1:32, 1:64, 1:128, 1:256, and 1:512) from participants for 1h at 37 °C. Then, the mix was added to the 96-well plates with the Vero E6 cells. Cytopathic effect on Vero E6 cells was analyzed 7 days after infection. For each test, a serum sample from uninfected patients (negative control) and a neutralizing COVID-19 patient serum sample (positive control) were used.

**Supplementary Figures**

**Figure S1. Titer evaluation of specific IgG anti-N of SARS-CoV-2 by ELISA assays in the serum of participants in the 18-59 years old and ≥60 years old, inoculated with CoronaVac or placebo in a 0-14 schedule.** OD_450_ obtained for the different dilution at (A) 0 days (1^st^ dose), (B) 14 days (2^nd^ dose), (C) 28 days after the first dose, and (D) 42 days after the first dose. Serial dilutions performed with the sera obtained from participants inoculated with CoronaVac (white symbols for participants in the 18-59 years old group and black symbols for participants in the ≥60 years old group) or placebo (grey symbols) were evaluated by ELISA assays in order to detect IgG specific against the N protein of SARS-CoV-2. Sera samples from a seropositive participant at the beginning of the study were included as positive controls (asterisk symbols), and from a seronegative participant as negative controls (cross circle symbols).

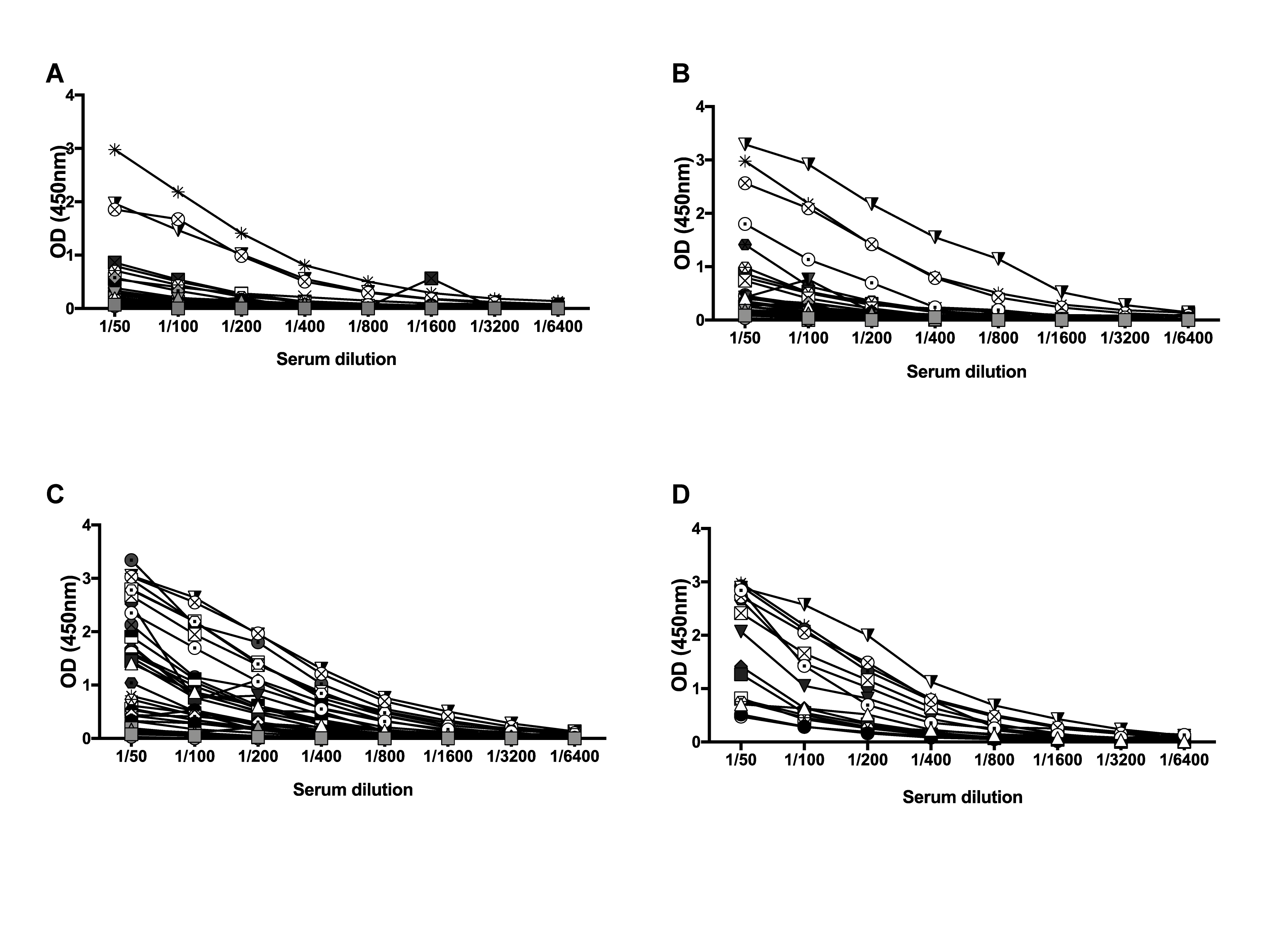

**Figure S2. Titer evaluation of specific IgG anti S1-RBD of SARS-CoV-2 by ELISA assays in the serum of participants in the 18-59 years old and ≥60 years old groups, inoculated with CoronaVac or placebo in a 0-14 schedule.** OD_450_ obtained for the different dilution at (A) 0 days (1^st^ dose), (B) 14 days (2^nd^ dose), (C) 28 days after the first dose, and (D) 42 days after the first dose. Serial dilutions performed with the sera obtained from participants inoculated with CoronaVac (white symbols for participants in the 18-59 years old group and black symbols for participants in the ≥60 years old group) or placebo (grey symbols) were evaluated by ELISA in order to detect IgG specific for S1-RBD of SARS-CoV-2. Sera samples from a seropositive participant at the beginning of the study were included as positive controls (asterisk symbols), and from a seronegative participant as negative controls (cross circle symbols).

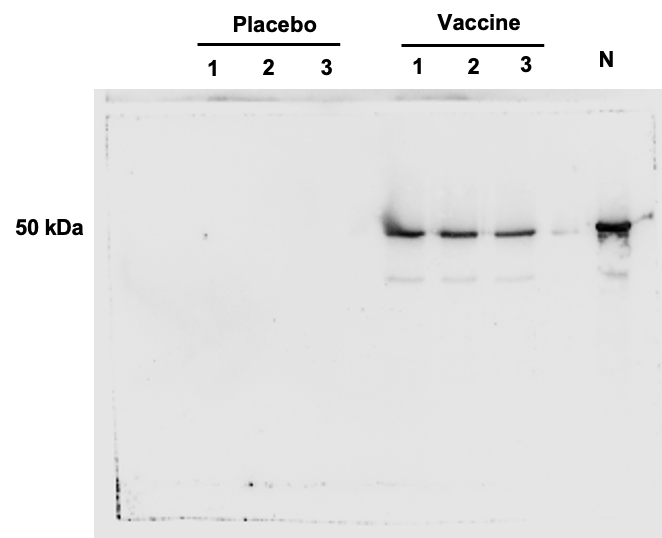

**Figure S3. Detection of SARS-CoV-2 N protein in samples of CoronaVac and placebo.** Samples of placebo and CoronaVac (225 ng each) were loaded in a 12% SDS-PAGE followed by western blot to detect SARS-CoV2-N protein. Recombinant N protein (1 μg, SinoBiological, 40588-V08B) was loaded as a positive control (Inverted N in gel, last lane). Membranes were incubated with a mouse anti-SARS-CoV-2 N antibody (1:1000, Sinobiological, 40143-MM05, Lot Nº HB14FE2110) followed by incubation with a secondary anti-mouse HRP-conjugated antibody (Invitrogen, 62-6520, Lot Nº UJ287702).

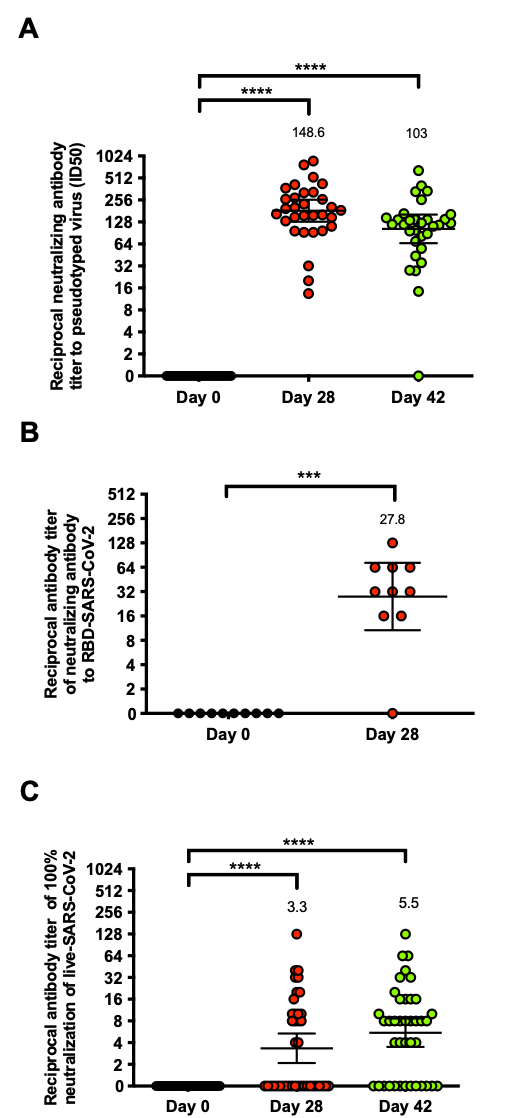

**Figure S4. Evaluation of the** **neutralizing capacity of antibodies against SARS-CoV-2 after two immunizations in a 0-14 schedule.** (A) Titers of neutralizing antibodies evaluated with a pseudotyped viral system ^15^. Data is represented as the reciprocal dilution of sera that prevent infection by 50% (ID_50_) after the first dose. Numbers above the bars show the Geometric Mean Titer (GMT) and the error bars indicate the 95% CI. (B) Titers of neutralizing antibody evaluated with a surrogate neutralization assay developed by BioHermes, evaluated at 0 and 28 days after the first dose. Data is represented as the reciprocal dilution of sera that inhibits over 10% the binding of HRP-RBD to the recombinant hACE2 protein after the first dose. Numbers above the bars show the Geometric Mean Titer (GMT) and the error bars indicate the 95% CI. Subjects from the vaccine arm included in the assays are 17 for A and 10 for B. (C) Neutralization assays in Vero E6 cells infected with live SARS-CoV-2 (33782CL-SARS-CoV-2 strain). Data is represented as the reciprocal dilution of sera that prevents cytopathic effect by 100% after the first dose. Numbers above the bars show the Geometric Mean Titer (GMT) and the error bars indicate the 95% CI. Data were analyzed with a one sample t test to assess if the mean is different to 1; ***p<0.0005, ****p<0.0001.

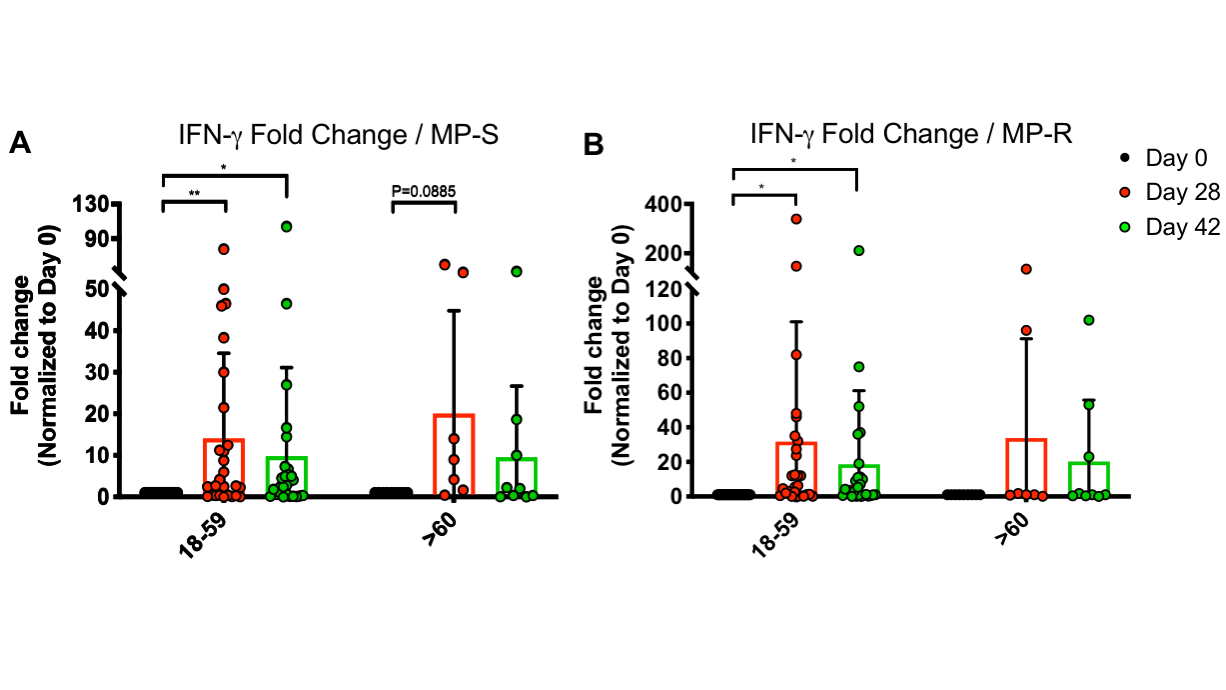

**Figure S5. Fold change of SFCs for IFN-γ measured by ELISPOT (MP-S and MP-R).** Changes in the secretion of IFN-γ, determined as the fold change normalized to Day 0, was measured. Data was obtained upon stimulation with an MP of peptides of the S-SARS-CoV-2 protein (A), and an MP of the remaining peptides of the virus (B). A total of 27 participants were included in the 18-59 age group and a total of 9 for the ≥60 years old group. Data were analyzed with a one sample t test to assess if the mean is different to 1; *p<0.05, **p<0.005.

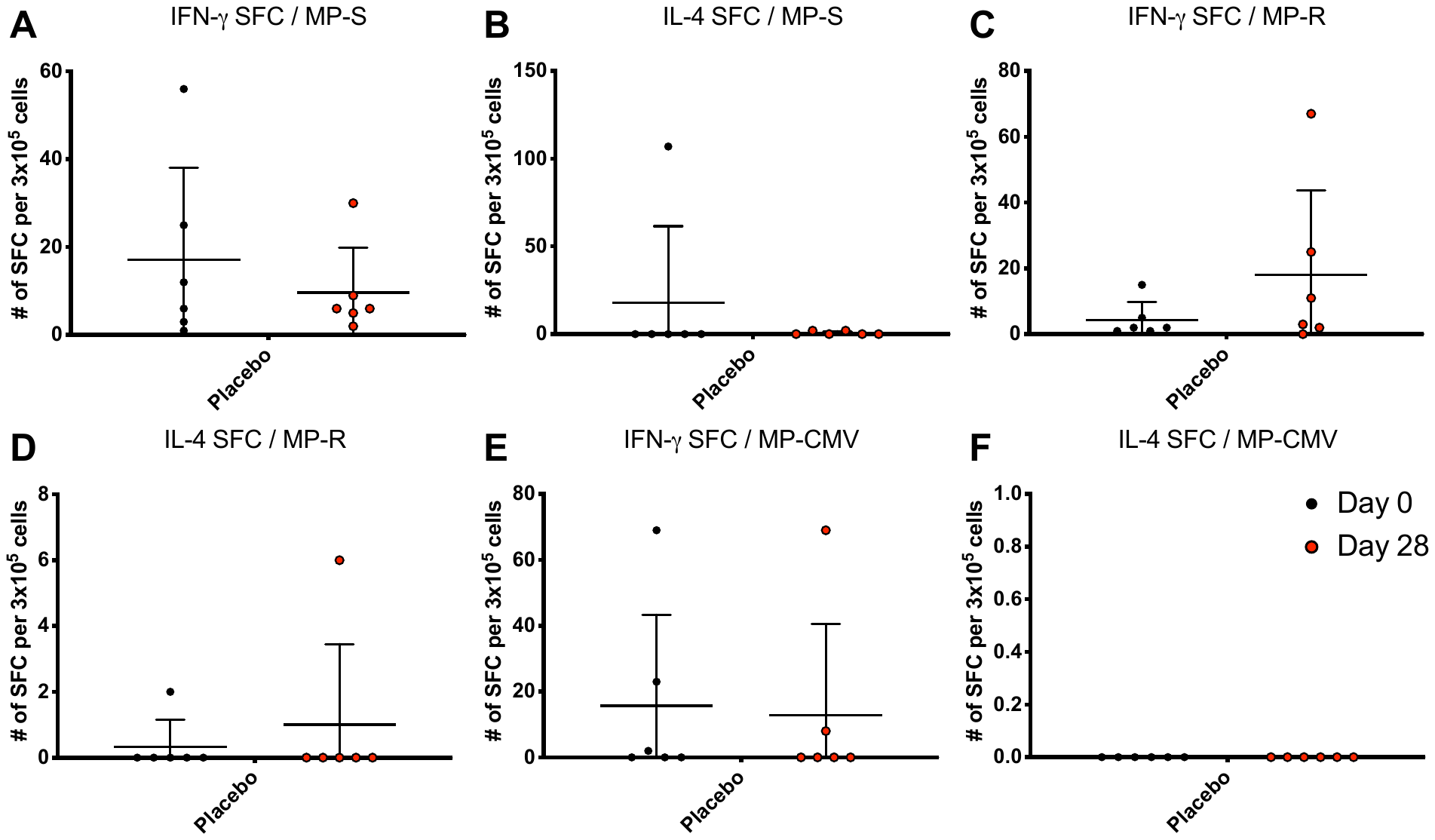

**Figure S6. Count of SFCs for IFN-γ and IL-4, as measured by ELISPOT, in the placebo group (MP-S, MP-R, and CMV).** The secretion of IFN-γ and IL-4, determined as the number of SFC, was measured and is shown upon stimulation with an MP of peptides of the S-SARS-CoV-2 protein (A-B), an MP of the remaining peptides of the virus (C-D), and an MP of peptides from CMV (E-F). 6 participants from the 18-59 years old group were included. Data from days 0 and 28 is shown, as no data for day 48 is available for these participants.

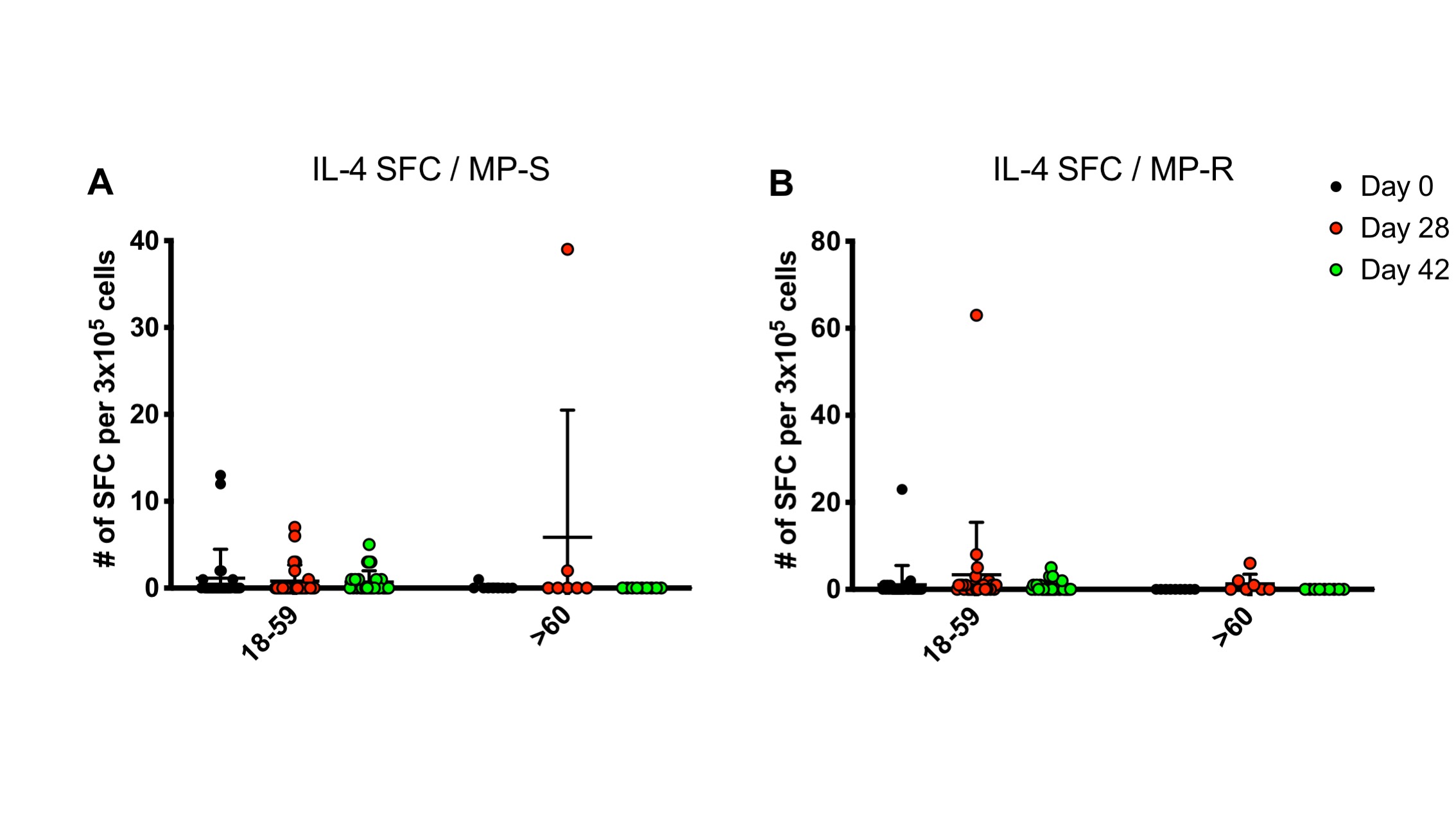

**Figure S7. Count of SFC for IL-4, as measured by ELISPOT (MP-S and MP-R).** The secretion of IL-4, determined as the number of SFCs, was measured and is shown upon stimulation with an MP of peptides of the S-SARS-CoV-2 protein (A), and an MP of the remaining peptides of the virus (B). A total of 27 participants were considered in the 18-59 age group and a total of 9 for the 60 years and older group. Data shown represent means ± SD. Data from each age group were analyzed separately by a Friedman test for repeated measures, followed by a *post hoc* Dunn’s test corrected for multiple comparisons against day 0 for each age group.

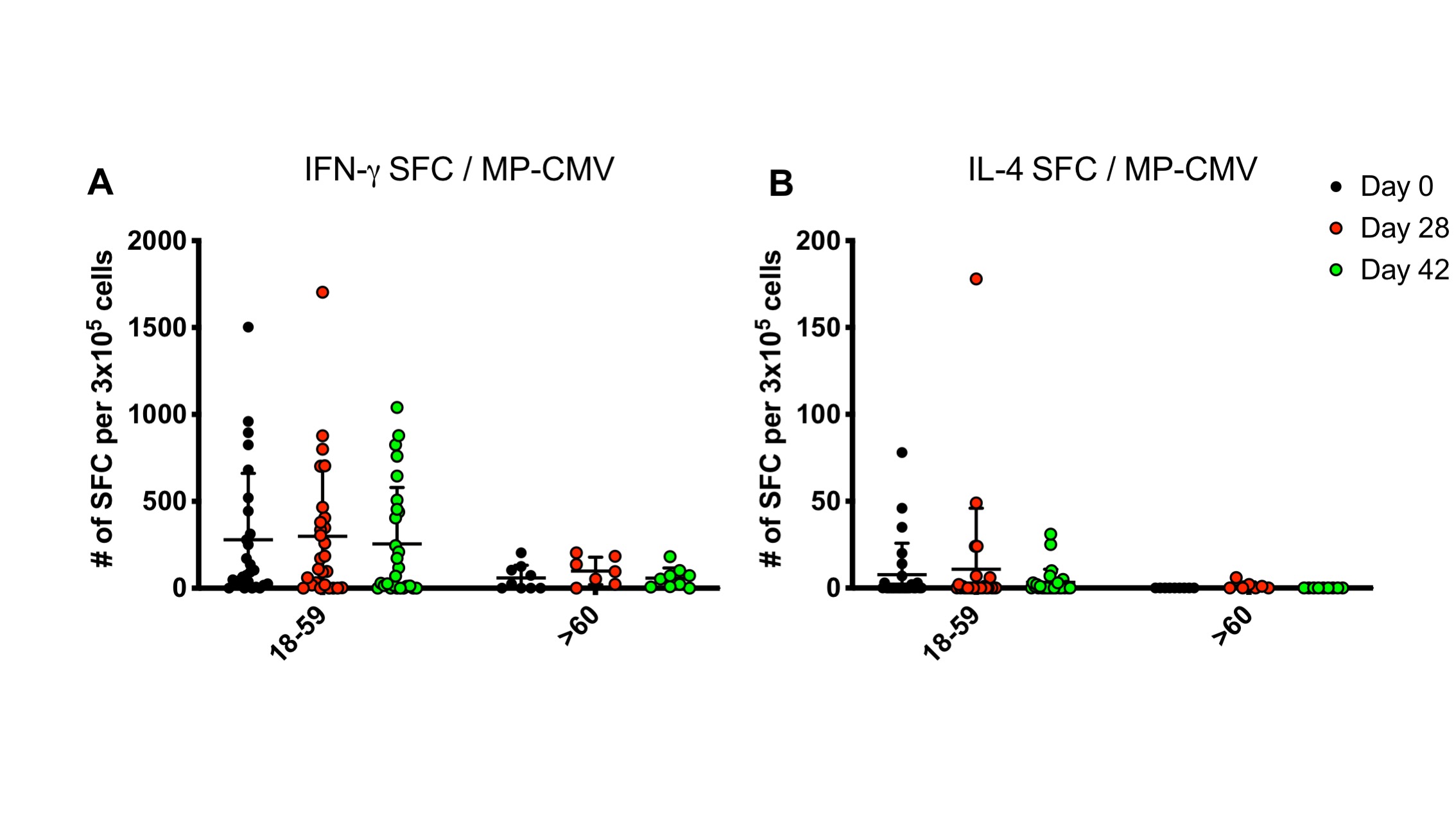

**Figure S8. Count of SFCs for IFN-γ and IL-4, upon stimulation with MP-CMV, as measured by ELISPOT.** The secretion of IFN-γ (A) and IL-4 (B), determined as the number of SFC, was measured and is shown upon stimulation with an MP of peptides of cytomegalovirus. A total of 27 participants were considered in the 18-59 years old group and a total of 9 for the ≥60 years old group. Data shown represent means ± SD. Data from each age group were analyzed separately by a Friedman test for repeated measures, followed by a *post hoc* Dunn’s test corrected for multiple comparisons against day 0 for each age group.

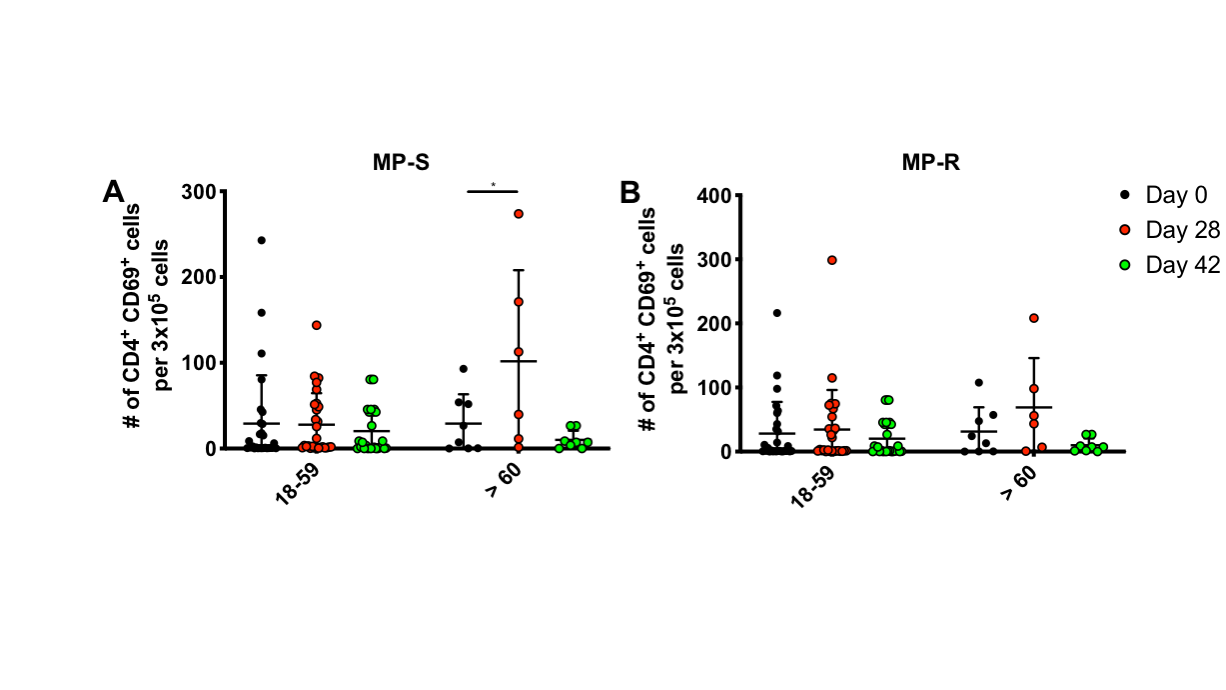

**Figure S9. Absolute count of activated CD4^+^ T cells upon stimulation with MP-S and MP-R, as determined by flow cytometry.** The number of activated CD4^+^ T cells (Defined as CD45^+^, CD3^+^, CD69^+^) was determined by flow cytometry, upon stimulation with a MP of peptides of the S-SARS-CoV-2 protein (A), and a MP of the remaining peptides of the virus (B). A total of 27 participants were considered in the 18-59 age group and a total of 9 for the 60 years and older group. Data from each age group were analyzed separately by a Wilcoxon test for repeated measures for comparisons against day 0 for each age group; *=p<0.05.

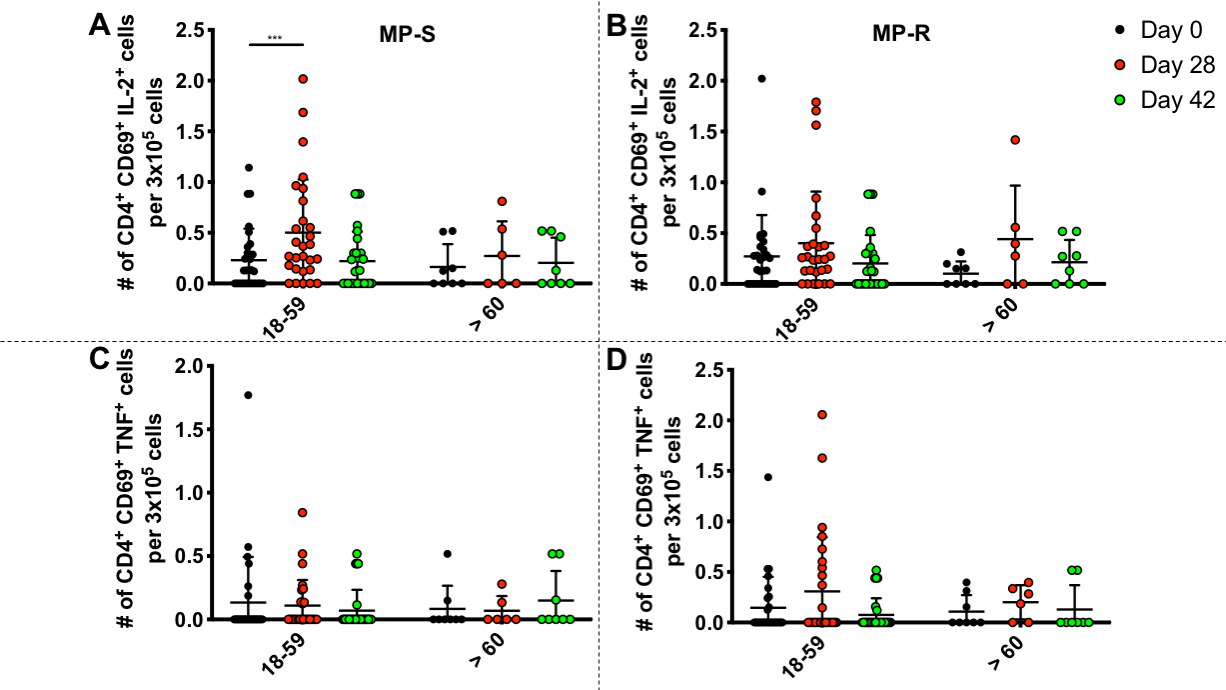

**Figure S10. Absolute count of activated CD4^+^ T cells secreting IL-2 or TNF-α upon stimulation with MP-S and MP-R, as determined by flow cytometry.** The number of activated CD4^+^ T cells (Defined as CD45^+^, CD3^+^, CD69^+^) secreting IL-2 (A, B) and TNF-α (C, D) were determined by flow cytometry, upon stimulation with an MP of peptides of the S-SARS-CoV-2 protein (left panels), and an MP of the remaining peptides of the virus (right panels). A total of 27 participants were considered in the 18-59 age group and a total of 9 for the 60 years and older group. Data from each age group were analyzed separately by a Wilcoxon test for repeated measures for comparisons against day 0 for each age group; ***p<0.0005.

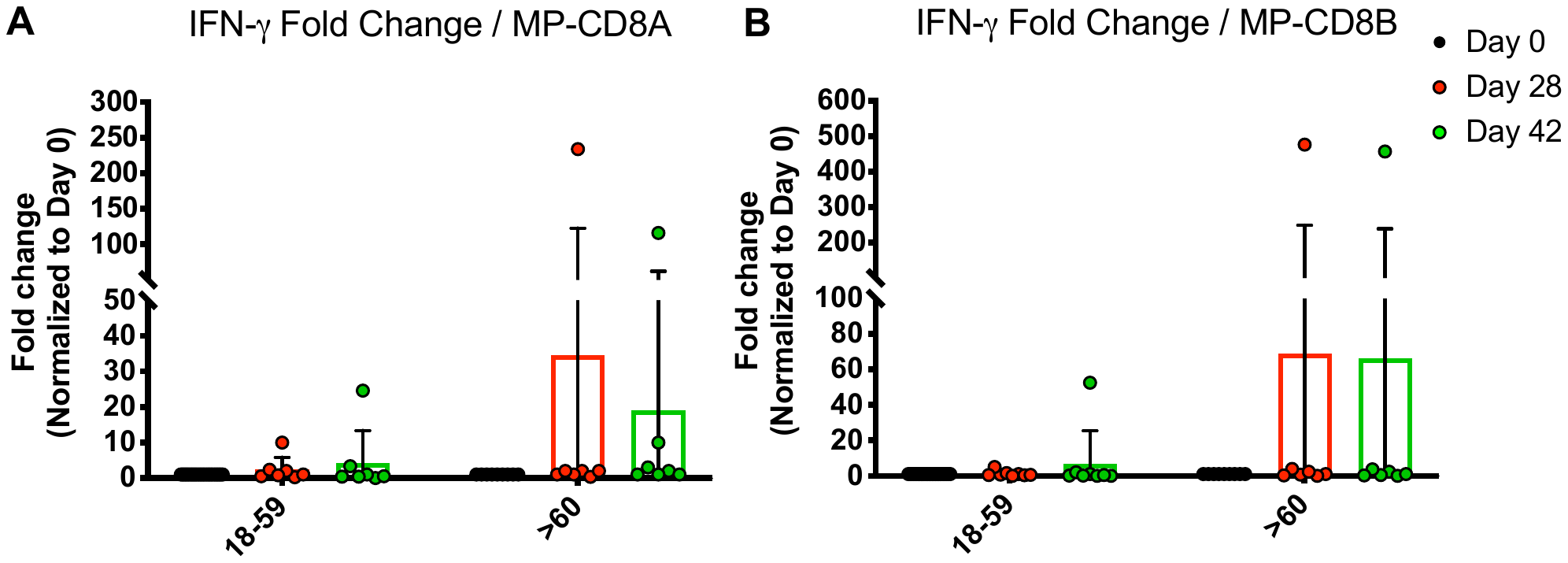
 **Figure S11. Fold change of SFCs for IFN-γ, as measured by ELISPOT upon stimulation with MP peptides (MP-CD8A and MP-CD8B).** Changes in the secretion of IFN-γ, determined as the fold change normalized to Day 0, was measured. Data was obtained upon stimulation with MP CD8A (A) and CD8B (B) of the S-SARS-CoV-2 protein. A total of 27 participants were included in the 18-59 age group and a total of 9 for the ≥60 years old group. Data were analyzed with a one sample t test to assess if the mean is different to 1.

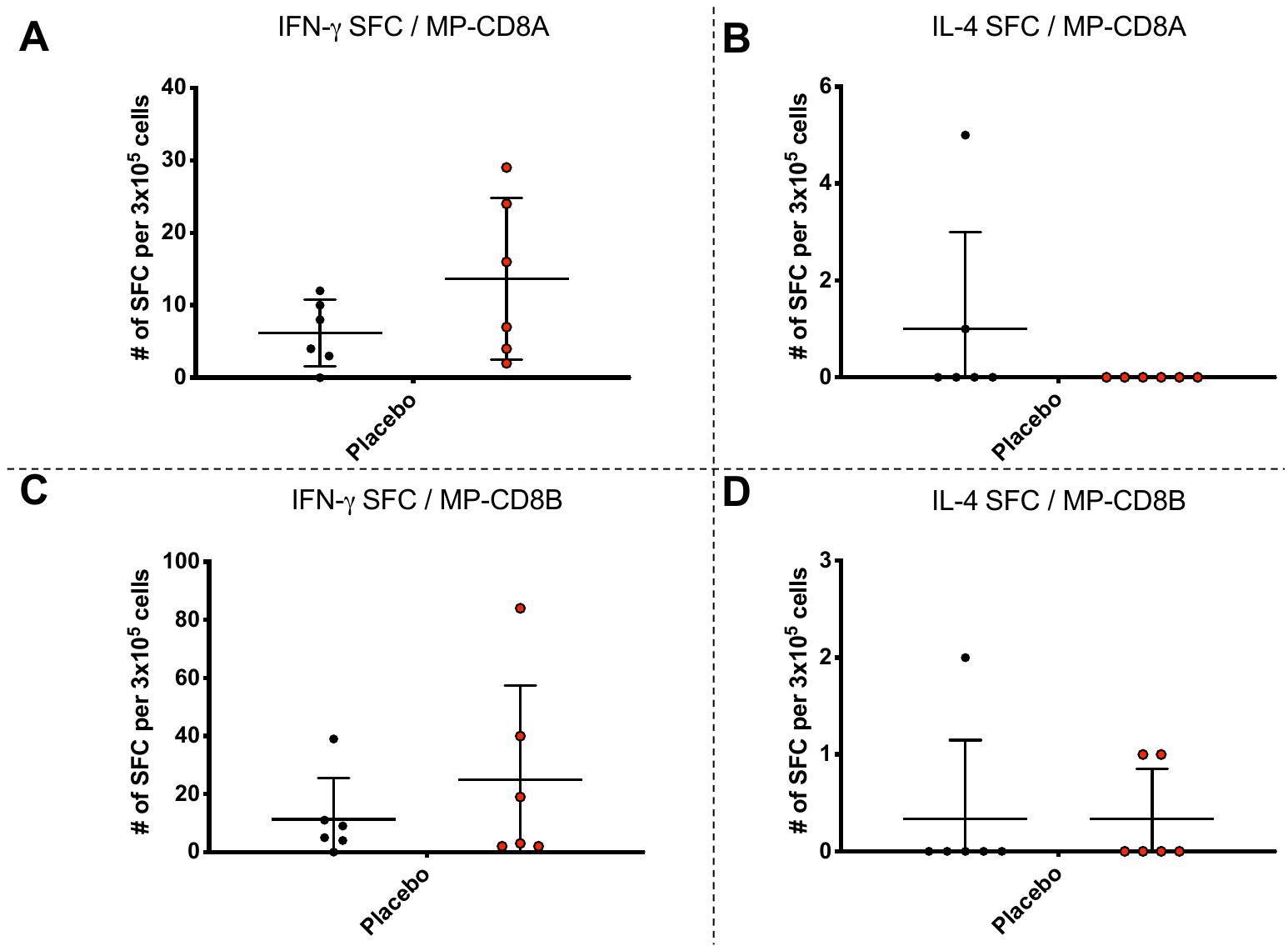
**Figure S12. Count of SFCs for IFN-γ and IL-4, as measured by ELISPOT, in the placebo group (MP-CD8A and MP-CD8B).** The secretion of IFN-γ and IL-4, determined as the number of SFC, was measured and is shown upon stimulation with MP of peptides of the SARS-CoV-2 proteins CD8A (A, C), and CD8B (B, D). A total of 6 participants from the 18-59 years old group from the placebo branch were included for this assay. Data from days 0 and 28 is shown, as no data for day 48 is available for these participants. Data were analyzed with a two-tailed paired Student’s t test; *p<0.05, **p<0.005, ***p<0.0005, ****p<0.0001.

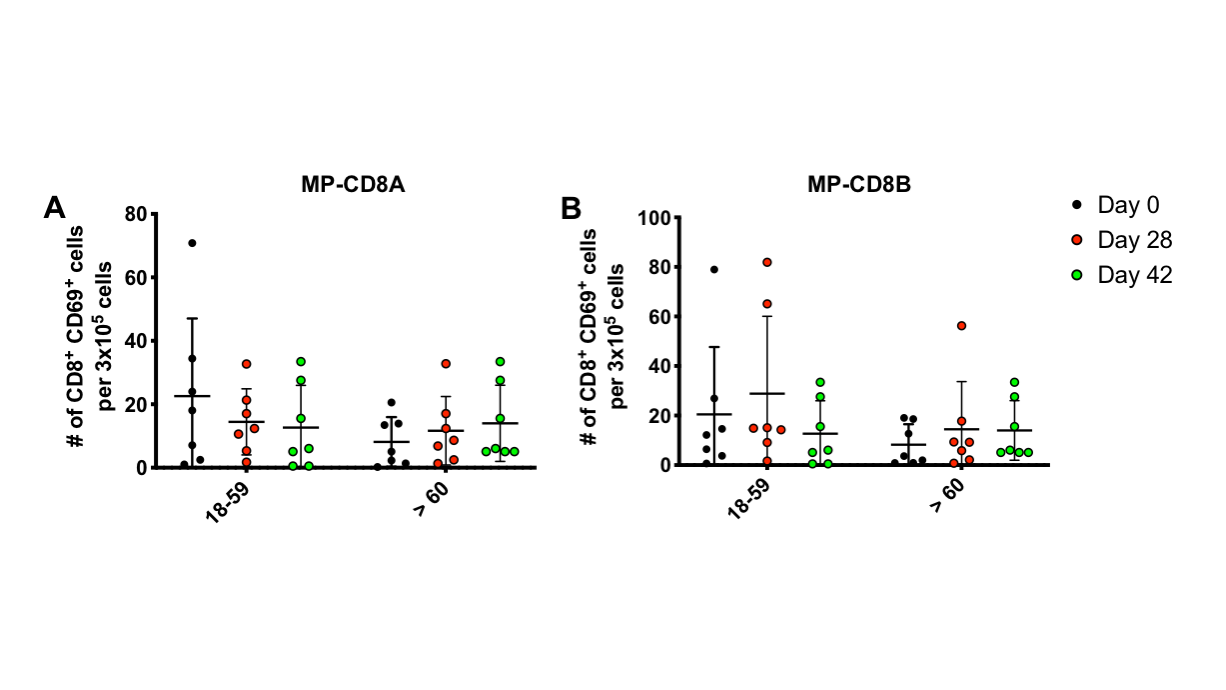

**Figure S13. Absolute count of activated CD8^+^ T cells upon stimulation with MP-CD8A and MP-CD8B, as determined by flow cytometry.** The number of activated CD8^+^ T cells (Defined as CD45^+^, CD3^+^, CD69^+^) was determined by flow cytometry, upon stimulation with two MP of peptides of SARS-CoV-2, designed to stimulate CD8^+^ T cells. A total of 7 participants were considered in the 18-59 age group and a total of 7 for the ≥60 years old group. Data from each age group were analyzed separately by a Wilcoxon test for repeated measures for comparisons against day 0 for each age group; *=p<0.05, **=p<0.005, ***p<0.0005, ****p<0.0001.

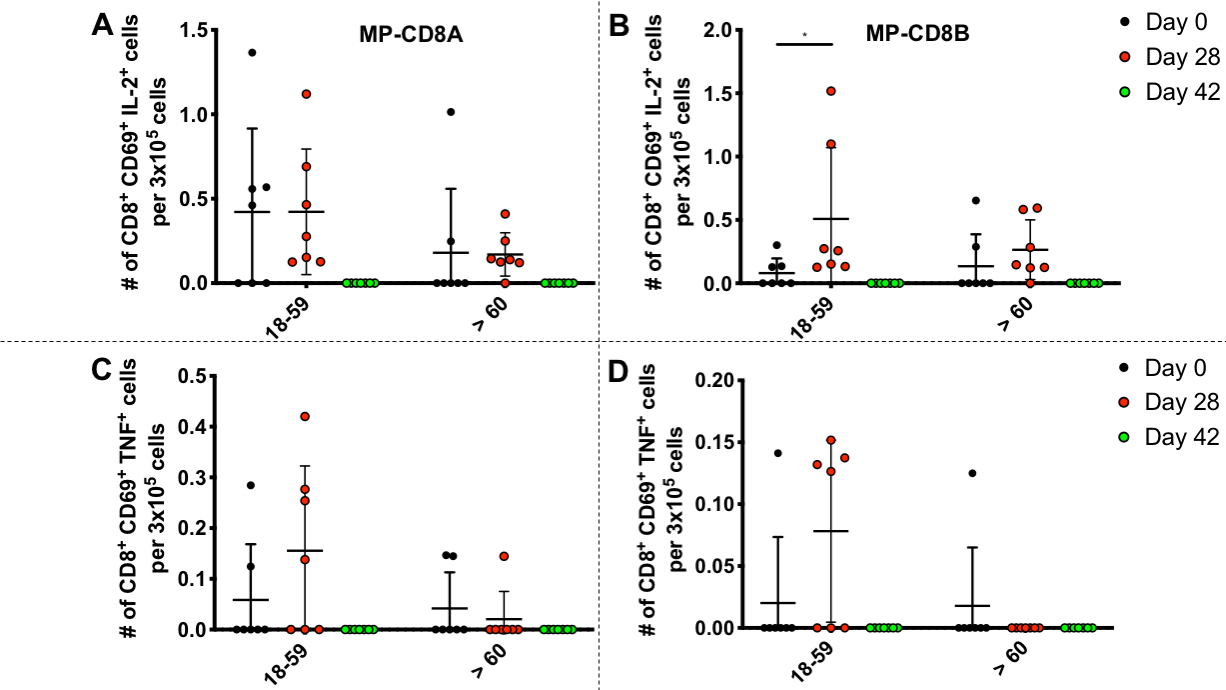

**Figure S14. Absolute count of activated CD4^+^ T cells secreting IL-2 or TNF-α upon stimulation with MP-CD8A and MP-CD8B, as determined by flow cytometry.** The number of activated CD4^+^ T cells (Defined as CD45^+^, CD3^+^, CD69^+^) secreting IL-2 (A, B) and TNF-α (C, D) were determined by flow cytometry, upon stimulation with MP of peptides of SARS-CoV-2 proteins CD8A (left panels), and CD8B (right panels). A total of 7 participants were considered in the 18-59 age group and a total of 7 for the ≥60 years old group. Data from each age group were analyzed separately by a Wilcoxon test for repeated measures for comparisons against day 0 for each age group; *=p<0.05.
