## Supplementary Annex for "Interim report: Safety and immunogenicity of an inactivated vaccine against SARS-CoV-2 in healthy chilean adults in a phase 3 clinical trial"

**Supplementary Appendix**

**1. CoronaVac03CL Study Team**

**Center CL1: Áreas Ambulatorias Marcoleta - Pontificia Universidad Católica de Chile.**

Alvaro Miguel Rojas Gonzalez, Maria Soledad Navarrete Bello, Constanza Belen Del Rio Solis, Dinely Valeska Del Pino Lavin, Natalia Elizabeth Aguirre Concha, Grecia Marly Salinas Escala, Franco Vega Farias, Acsa Raquel Salgado Ovalle, Thomas Quinteros, Marlene Ortiz, Marcela Puente, Alma Muñoz, Patricio Astudillo, Monique Nicole Le Corre.

**Center CL2: Clínica San Carlos de Apoquindo - Red de Salud UC-Christus**

Marcela Potin Santander, Juan Catalán Henríquez, Melan Peralta Kong, Consuelo Zamanillo Moreira, Nicole Keller Riquelme, Rocio Fernández Bustos, Sofia Aljaro Ehrenberg, Sofia López Colomba, José Tomas González, Tania Weil Valjalo, Luz Opazo Barrientos, Paula Muñoz Galeb, Inés Estay Puebla, Miguel Cantillana Morales, Liliana Carrera del Canto, Matías Masalleras Oyarzún.

**Center CL4: Clínica Los Andes - Universidad de Los Andes**

Paula Guzmán Merino, Francisca Aguirre Boza, Aarón Cortés Rojas, Luis Federico Bátiz, Javiera Francisca Pérez Velásquez, Karen Pamela Apablaza García, Lorena Yates Barsotti, María de los Ángeles Valdés Valdés, Bernardita Hurtado, Veronique Venteneul, Constanza Astorga.

**Center CL5: Clínica Alemana - Universidad del Desarrollo**

Paula Andrea Muñoz-Venturelli, Pablo Agustín Vial, Andrea Ingrid Schilling Redlich, Daniela Pavez Azurmendi, Inia Andrea Pérez Villa, Amy Lisa Riviotta, Francisca Gonzalez Mc Cowley, Francisca Pilar Urrutia Goldsack, Alejandra Isabel Del Río Weldt, Claudia Andrea del Carmen Asenjo Lobos, Bárbara Paulina Vargas Latorre, Francisca Valentina Castro Fuentes, Alejandra Patricia Acuña Rogel, Javiera Constanza Gúzman Cancino, Camila Alejandra Astudillo Griffiths.

**Center CL6: Hospital Clínico Félix Bulnes - Universidad San Sebastián**

Carlos M Pérez, Pilar Espinoza, Andrea Martínez, Marcela Arancibia, Harold Romero, Cecilia Bustamante, María Loreto Pérez, Natalia Uribe, Viviana Silva, Bernardita Morice, Marco Pérez.

**Center CL7: Hospital Dr. Gustavo Fricke - Universidad de Valparaíso**

Marcela González, Werner Jensen, Claudia Pasten, Ma. Fernanda Aguilera, Nataly Martínez, Camila Molina, Sebastián Arrieta, Begoña López, Claudia Ortiz, Macarena Escobar, Camila Bustamante, Marcia Espinoza, Angela Pardo, Alison Carrasco, Miguel Montes, Macarena Saldías, Natalia Gutiérrez, Juliette Sánchez.

**Center CL8: Hospital Carlos Van Buren- Universidad de Valparaíso**

Daniela Fuentes Hulse, Yolanda Calvo Toro, Mariela Cepeda Corrales, Rosario Lemus Manzur, Muriel Suarez Saavedra, Mercedes Armijo Rodríguez, Shirley Monsalves González, Constance Marucich Baeza, Cecilia Cornejo Beas, Ángela Acosta Palacio, Xaviera Prado, Francisca Yáñez, Marisol Barroeta Andrade, Claudia López García.

**Center CL9: Complejo Asistencial Dr. Sótero del Rio**

Paulina Donato Inostroza, Martin Lasso Barreto, María Iturrieta Meléndez, Juan Giraldo Paramo, Francisco Gutiérrez Valenzuela, María Acuña Schlegel, Ada Cascone Scarpati, Raymundo Rojas Araya, Camila Sepúlveda Contreras, Mario Alex Contreras, Yessica Campisto Sanhueza, Pablo González Sanhueza, Zoila Quizhpi Mejias, Mariella Lopez García, Vania Pizzeghello Salfate, Stephannie Silva Monsalve.

**2. Members of the Independent Data Safety Monitoring Committee.**

Luis Delpiano, MD, Pediatric Infectologist, Hospital San Borja Arriarán, Santiago, Chile.

Macarena Lagos, MD, Immunologist, Clínica Las Condes and Hospital Padre Hurtado, Santiago, Chile.

Gloria Icaza, MD, Epidemiologist and Statitician, Universidad de Talca, Talca, Chile.

Leonardo Chanqueo, MD, Adult Infectologist, Hospital San Juan de Dios, Santiago, Chile.

Mónica Imarai, PhD, Universidad de Santiago, Santiago, Chile.
